# Optimizing hospital opioid deprescribing: a multi-level consensus study bridging evidence, expertise, and patient perspectives

**DOI:** 10.1101/2025.05.27.25328396

**Authors:** Marcel Rainer, Elin Sebesta, Maria Monika Wertli, Andrea Michelle Burden, Dominik Stämpfli

**Author notes:** Corresponding author: Dominik Stämpfli, Institute of Pharmaceutical Sciences, ETH Zurich, Vladimir-Prelog Weg 1-5/10, 8093 Zurich, Switzerland.

## Abstract

Hospital-initiated opioid analgesics that extend beyond discharge can lead to long-term use and adverse outcomes. Despite a growing understanding of opioid-related harms, there is a lack of published protocols for structured deprescribing in Europe. We aimed to develop an actionable opioid deprescribing framework that builds on trialed reduction protocols with patient-centered determinants for systematic implementation in tertiary care. We conducted a multi-level consensus study to bridge evidence, clinician expertise, and patient perspectives. Initial framework development included focus group discussions with multidisciplinary clinicians (n=5). The framework was validated for our institution via a two-round Delphi survey across six medical specialties (n=11). An opioid reduction calculator was developed in Python (version 3.12.3), incorporating clinical parameters to determine reduction trajectories for prespecified starting and end doses. Final refinement included interviews with patients receiving opioid therapy (n=11) to optimize understandability of the reduction plan as a patient handout and for developing a patient pamphlet on opioids with patients (n=13). The framework identified four critical domains for successful opioid deprescribing: interventional reduction strategies, patient-specific physiological variables, environmental enablers, and procedural elements. Reduction strategies categorized patients into chronic and new users, incorporating grace periods for dose stabilization, with particular attention given to patients exhibiting pain catastrophizing behavior and frailty. Environmental and procedural factors included shared responsibility and interdisciplinarity, ensuring that follow-up is facilitated by a coordinated care team. These findings were operationalized into the reduction calculator, proposing initial individualized reduction plans (10-25%) based on key patient-specific variables with built-in stabilization periods. Delphi validation achieved full consensus (87.8% first-round agreement; 100% final consensus) on all framework components. Patient involvement refined the handout to improve understandability and actionability, and yielded comprehensive information on opioids. We developed an actionable opioid deprescribing framework that bridges published evidence with patient-centered care for our institution, providing other healthcare institutions with a practical blueprint for their own implementation. Our multi-level consensus identified critical patient-specific and environmental determinants, and established individualized reduction protocols, addressing a critical gap in transitional care while facilitating safe opioid deprescribing practices.

## Background

Opioid analgesics are frequently initiated in hospitals to manage acute pain from surgery or trauma (1). However, these initial opioid prescriptions are commonly continued after discharge, even after acute pain subsides, resulting in prolonged opioid use in some patients (2,3). Hofer et al. (4) analyzed European postoperative opioid use trajectories and found that approximately 1% of opioid-naïve patients remained on opioids over a year after the initial postoperative period, transitioning from acute to long-term use. These patients reported higher pain levels and greater interference with daily activities, with 1 in 10 having developed chronic pain unrelated to the surgery responsible for the first opioid prescription. General practitioners (GPs), who frequently assume responsibility for these prescriptions, may face challenges in determining appropriate reduction protocols due to limited guidance (5,6). Such findings underscore the need for structured approaches to prevent long-term opioid use after hospitalization.

In response, opioid stewardship initiatives have been developed to monitor opioid use, optimize prescribing practices, reduce long-term use, and prevent opioid-related adverse effects. Opioid stewardship is defined as a multidisciplinary, coordinated, systematic approach to opioid prescribing that emphasizes safe, effective pain management while minimizing the potential for misuse and dependence (7). As a strategy to address opioid overuse, opioid stewardship initiatives can implement individualized opioid reduction protocols, particularly after acute care episodes. Such opioid exit plans (OEPs) can substantially reduce post-acute opioid use, with individualization as an integral part (8) of the deprescribing framework. For example, Hah and colleagues (9) reported a faster return to baseline opioid use and a 62% higher rate of successful discontinuation (inverted hazard ratio=1.62, p=0.03) in the intervention group compared to standard care.

Despite growing documented local associations between opioid prescribing and increased odds of rehospitalization (10,11) and emergency department visits (12,13), Europe currently lacks published opioid stewardship initiatives, such as OEPs. In addition, a recent review of opioid deprescribing guidelines found that practical recommendations are still mostly missing (14). To address this unmet need for standardized protocols for opioid deprescribing, we conducted a multi-level consensus study that merged expertise from clinicians and patient perspectives with current guidelines and international initiatives to construct an actionable and locally accepted opioid deprescribing framework to serve as a rationale for OEPs. This study aimed to identify patient-specific and environmental deprescribing determinants, opioid reduction protocols, and comprehensible patient information for potential adoption in clinical practice.

## Methods

A multi-level consensus approach was employed to develop an opioid deprescribing framework to reduce opioid analgesics in hospitalized patients. This consisted of deprescribing determinants, an opioid reduction calculator, a reduction plan handout, and a patient pamphlet on opioid use in general. Figure 1 outlines the study flow and development process. The overarching framework for opioid deprescribing consisted of two phases to incorporate both clinician and patient input. First, semi-structured focus group discussions were used to translate current evidence on pain management and opioid prescribing into deprescribing determinants and an opioid reduction calculator. These recommendations were then refined and brought to consensus through a Delphi survey with additional clinicians from a broader range of medical specialties. Following the initial validation phase, a reduction plan handout and patient pamphlet were generated from the revised recommendations and opioid calculator outputs. The final validation phase incorporated patient involvement to evaluate the developed reduction plan and patient pamphlet in structured interviews.

**Figure 1.**
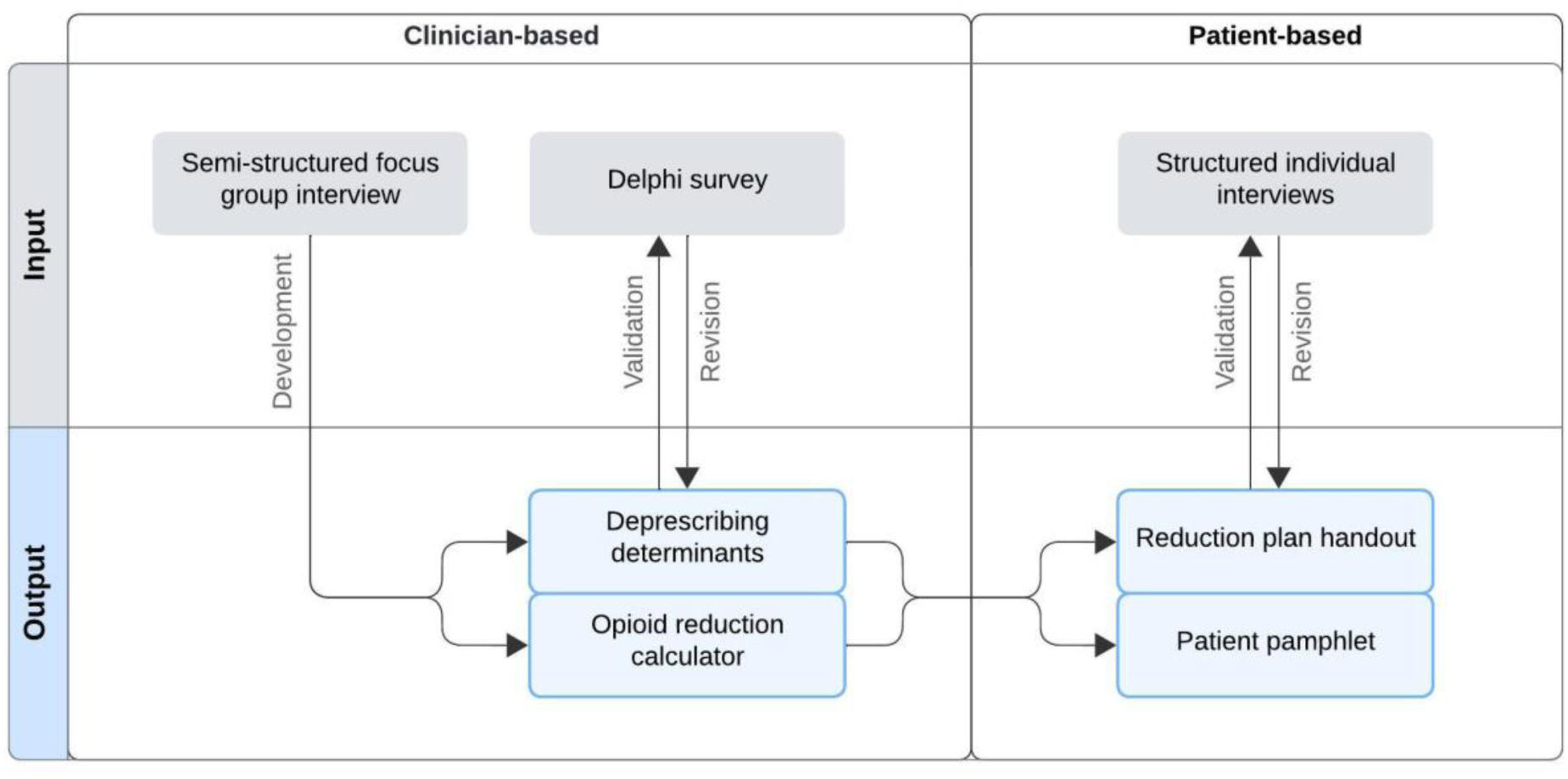
Study flow of the multi-level consensus approach to develop an actionable and locally accepted opioid deprescribing framework. The methodology employed an iterative approach to framework development. Initially, semi-structured focus group discussions were conducted to translate evidence-based pain management principles into deprescribing determinants and to develop an opioid reduction calculator. These preliminary determinants were validated through a Delphi survey involving medical specialists across multiple disciplines. Following primary validation, we developed patient materials, including reduction plan handouts and a patient pamphlet. The final phase incorporated structured patient interviews to evaluate comprehensibility and actionability of these materials, ensuring the framework had maintained patient-centeredness.

### Setting, data collection, and interim steps

A semi-structured interview guide was developed to facilitate the focus group discussion with clinicians. This guide incorporated a systematic review of OEPs (8), clinical guidelines of opioid reduction (15–17), and already existing internal institutional protocols. Eligible participants held a medical license to practice in Switzerland, had multiple years of experience managing pain with opioid analgesics, and represented different medical specialties. We recruited participants through purposive and snowball sampling within our hospital.

The interview was opened with a patient case followed by sequential questions focusing on patient-related, medication-related, and institutional factors. Participants were asked to discuss both recommendations of opioid reduction guidelines (15–18) and trialed OEPs (19–23) in general and in relation to the patient case presented. The interview took place in our hospital for 90 minutes and was audio-recorded for data collection. Details of the focus group setting and the interview guide are included in the supplement (section 1).

An opioid reduction calculator was developed based on the focus group findings that addressed the individualization of the reduction dosing. The calculator allows users to enter current opioid analgesics and dosages, specify target opioids and doses, and generate a reduction plan using conversion factors from the American Centers for Disease Control (15) and the identified stepwise reduction strategies. The calculator was implemented in Python (version 3.12.3) within the Anaconda (version 24.1.2) distribution and Visual Studio Code (version 1.91.1) environment that effectively combines morphine equivalents and the interviewees proposed reduction rates to suggest daily dosages for various opioids. The Python script of the calculator with its architecture are included in the supplement (sections 2.2 and 2.3).

The focus group recommendations and the calculator were evaluated and validated through a two-step Delphi survey to reach consensus and acceptance among clinicians from a wider range of medical specialties that prescribe opioids. Eligible participants were clinicians from medical specialties who were identified as top opioid prescribers in our hospital. We recruited participants through purposive and snowball sampling. Participants were emailed 49 survey questions in an Excel spreadsheet, clustered to represent the themes of the focus group findings. Participants were asked to rate on a 7-point Likert scale and to return the completed survey by email to a researcher or upload it to a cloud service provided by ETH Zurich. Items and recommendations that did not reach consensus in the first round were reworded, incorporating feedback from the respondents. In the second round, participants received the reworded items without previous ratings and unchanged items with anonymized pooled ratings as means and 95% confidence intervals (CIs). Participants were reminded once by email and once by phone to submit their responses and were marked as dropouts if they did not participate. The details of the Delphi survey are included in the supplement (section 3).

A reduction plan handout and patient pamphlet were developed based on the results from the validated focus group recommendations and a sample Swiss-German opioid pamphlet published on “Prevention with Evidence in Practice” (PEPra), a Swiss information platform to promote prevention and early detection established by the Swiss Medical Association and other supporting organizations (27). Following the recommendations on comprehensible patient information leaflets by Lampert and colleagues (28), the written contents’ readability was improved in incremental steps that could be verified using the Flesch Reading Ease Score (29). Subsequently, drafts of the reduction plan handout and the patient pamphlet were discussed in one-on-one patient interviews to incorporate patient perspectives. Patients were recruited at our hospital by two researchers (MR, DS) and were eligible for the interviews if they received opioid analgesics for any indication. All patients were asked 13 questions, consisting of three questions testing the patients’ understanding, time to read, structure questions, and sections to change. Each interviewer noted nonverbal expressions. Additionally, patients were asked to rate the overall comprehensibility on a 5-point scale (1 = not understandable, 5 = very understandable) and if they were to recommend the material to other patients. Feedback from these interviews was used to refine the drafts to ensure clarity, usability, and a patient-centered design. The interview questions are included in the supplement (section 4).

### Data analysis

For the focus group, the discussion was transcribed verbatim and contributions were anonymized in the process. We performed a deductive content analysis using a coding tree based on the clustering of the Consolidated Framework for Implementation Research (CFIR) 2.0 (30), with adaptations for local contexts and our research question. The coding tree was clustered into four domains following the CFIR methodology, and respective subcategories (major themes, subthemes) were iteratively refined through inductive content analysis, as detailed in the supplement (sections 1.2 and 1.3). Two researchers (MR, ES) independently coded interview transcripts using MAXQDA (version 4.3) (31). Discrepancies were resolved through in-person discussions to maintain inter-rater reliability, quantified as intercoder agreement and the Brennan-Prodiger coefficient (32). We performed systematic text condensation to formulate deprescribing recommendations and interaction analysis to identify thematic relationships (33,34). Participants received the researchers’ conclusions in a Word file and were invited to review the content. The focus group findings were manually clustered into themes and synthesized to create the overarching framework for opioid deprescribing with actionable recommendations.

For the Delphi survey, consensus was defined as ≥80% per item and if recommendations were rated ≥4.5 (“partially agree”) on a 7-point Likert scale. Respondents’ comments on individual items were incorporated into the content analysis of the focus group. Stability was assessed among the average absolute rating using a test-retest reliability coefficient of ≥0.70 (35). Statistical analysis was performed in RStudio (version 4.4.1) in the R environment (36,37). Packages psych (version 2.4.6.26) and tidyverse (version 2.0.0) were used (38,39).

For the patient interviews, we calculated the overall improvement in readability of the adapted information and the reported overall comprehensibility. We used content analysis to analyze the interview responses.

### Ethics, funding, and reporting

Ethical approvals were sought from the ETH Zurich Ethics Commission (focus group: EK-2024-N-37; patient interviews: Project 24 ETHICS-270). All study participants provided written informed consent. Financial compensation was provided to focus group and Delphi survey participants. No compensation was provided to structured interview participants. The study was funded by a research grant provided by the Swiss Association of Public Health Administration and Hospital Pharmacists (GSASA, “Forschungsprojekt nationaler Tragweite 2023”) (40). The study is reported in accordance with the consolidated criteria for reporting qualitative research (COREQ) (41) and reporting guidelines for mixed-methods research in health services (42). The checklists are included in the supplement (section 5).

## Results

### Focus group discussion

A total of five participants were recruited from different hierarchal levels, years of professional experience, and medical specialties. The characteristics are detailed in Table 1.

**Table 1.** Demographic and professional characteristics of the participants involved in the focus group discussion and Delphi survey.

| Characteristic | Focus Group Discussion (n=5) | Delphi Survey (n=13) |
| --- | --- | --- |
| **Age (years)** |  |  |
| Mean (Standard Deviation) | 41.2 (9.9) | 42.2 (8.9) |
| Range (Median) | 29-52 (43) | 29-52 (45) |
| **Gender** |  |  |
| Female, n (%) | 3 (60.0%) | 5 (38.5%) |
| Non-binary, n (%) | - | 1 (7.7%) |
| **Hierarchical Level** |  |  |
| Junior Physicians, n (%) | 2 (40.0%) | 2 (15.4%) |
| Senior Physicians, n (%) | 2 (40.0%) | 10 (76.9%) |
| Chief Physician, n (%) | 1 (20.0%) | 1 (7.7%) |
| **Years of Professional Experience** |  |  |
| Mean (Standard Deviation) | 13.8 (9.8) | 16.0 (8.8) |
| Range (Median) | 3-26 (14) | 3-26 (18) |
| **Medical Specialty** |  |  |
| Internal Medicine, n (%) | 2 (40.0%) | 4 (30.7%) |
| Orthopedics/Traumatology, n (%) | 2 (40.0%) | 1 (7.7%) |
| Pain Medicine, n (%) | 1 (20.0%) | 2 (15.4%) |
| Rheumatology, n (%) | - | 2 (15.4%) |
| Visceral Surgery, n (%) | - | 2 (15.4%) |
| Neurosurgery, n (%) | - | 2 (15.4%) |

Contributions to the discussion varied largely per participant from 7.5% to 38.8%, but were equally distributed among medical specialties with 31% coming from internal medicine, 30% coming from orthopedics/traumatology, and 39% coming from pain medicine. Following the coding tree, the findings were clustered into four domains (opioid reduction strategies, patient-specific variables, environmental enablers, and procedural factors) and translated into 49 recommendations, with six major themes and 15 subthemes identified. A detailed description of these subcategories is provided in the supplement (section 1.2). After determining the coding units, the intercoder agreement was 90.82% and the Brennan-Prodiger coefficient was determined to be 0.90 after face-to-face discussions.

### Delphi survey

Thirteen Delphi participants evaluated and validated the focus group findings, with two dropouts. Participants represented different hierarchal levels, years of professional experience, and medical specialties. The characteristics are detailed in Table 1 as well.

In the first round, consensus was achieved for 43 of the 49 items and recommendations (87.8%). The six items that did not reach consensus were revised for the second round. In the second round, all items achieved over 80% consensus. Reliability testing for unchanged items returned an average inter-rater reliability of 0.84 (95%-CI 0.77-0.89, p<0.001) between the first and second rounds, indicating sufficient stability (≥0.70) of the responses to close the Delphi survey. Non-consensus items and their revisions are included in the supplement (sections 3.2 and 3.3).

### Deprescribing determinants

Four key domains emerged from the content analysis of the focus group discussion and Delphi survey findings needed for successful opioid dose reduction: (1) opioid reduction strategies, (2) patient-specific variables, (3) environmental enablers, and (4) procedural factors. Table 2 summarizes the identified domains with their respective synthesized recommendations.

**Table 2.**
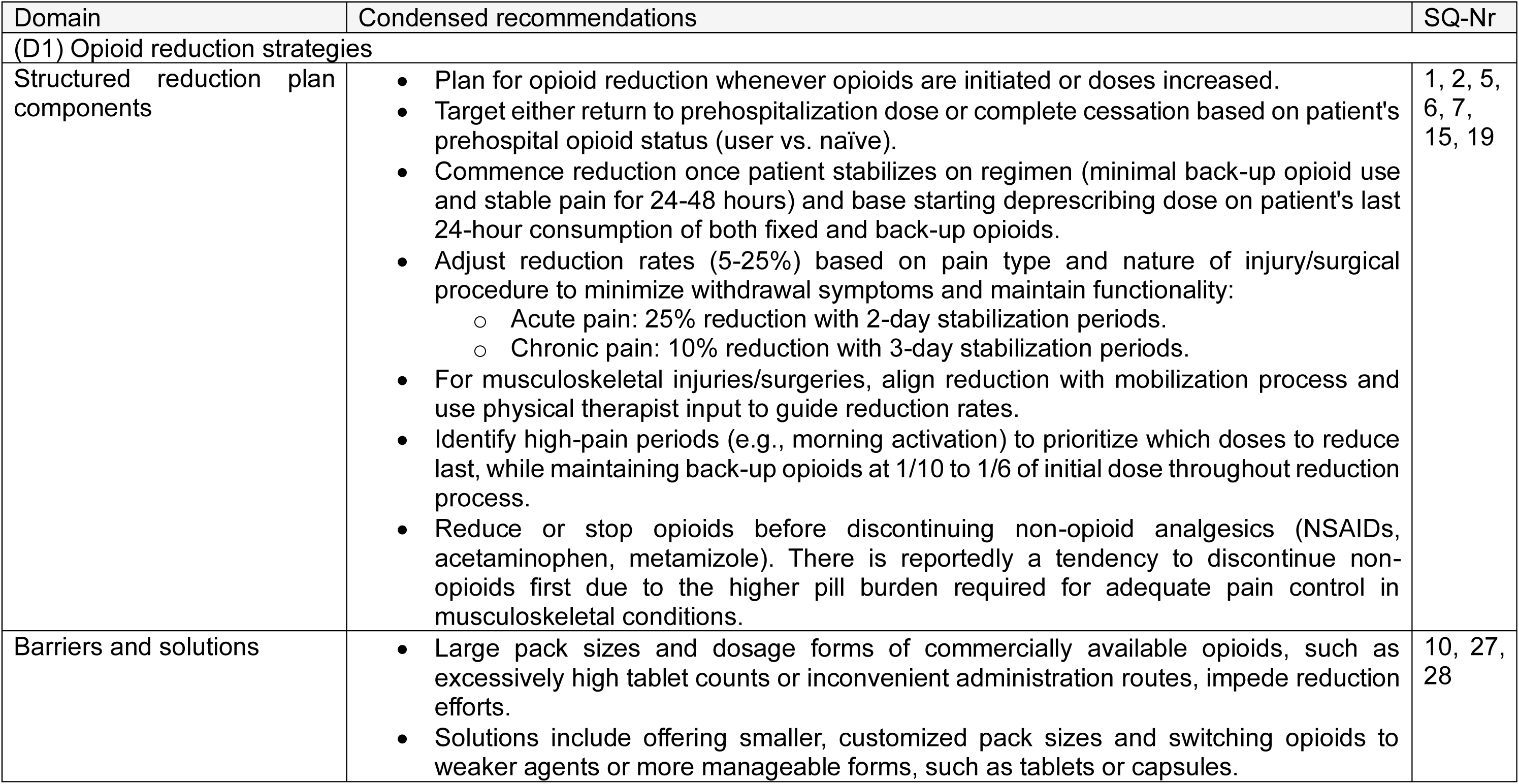
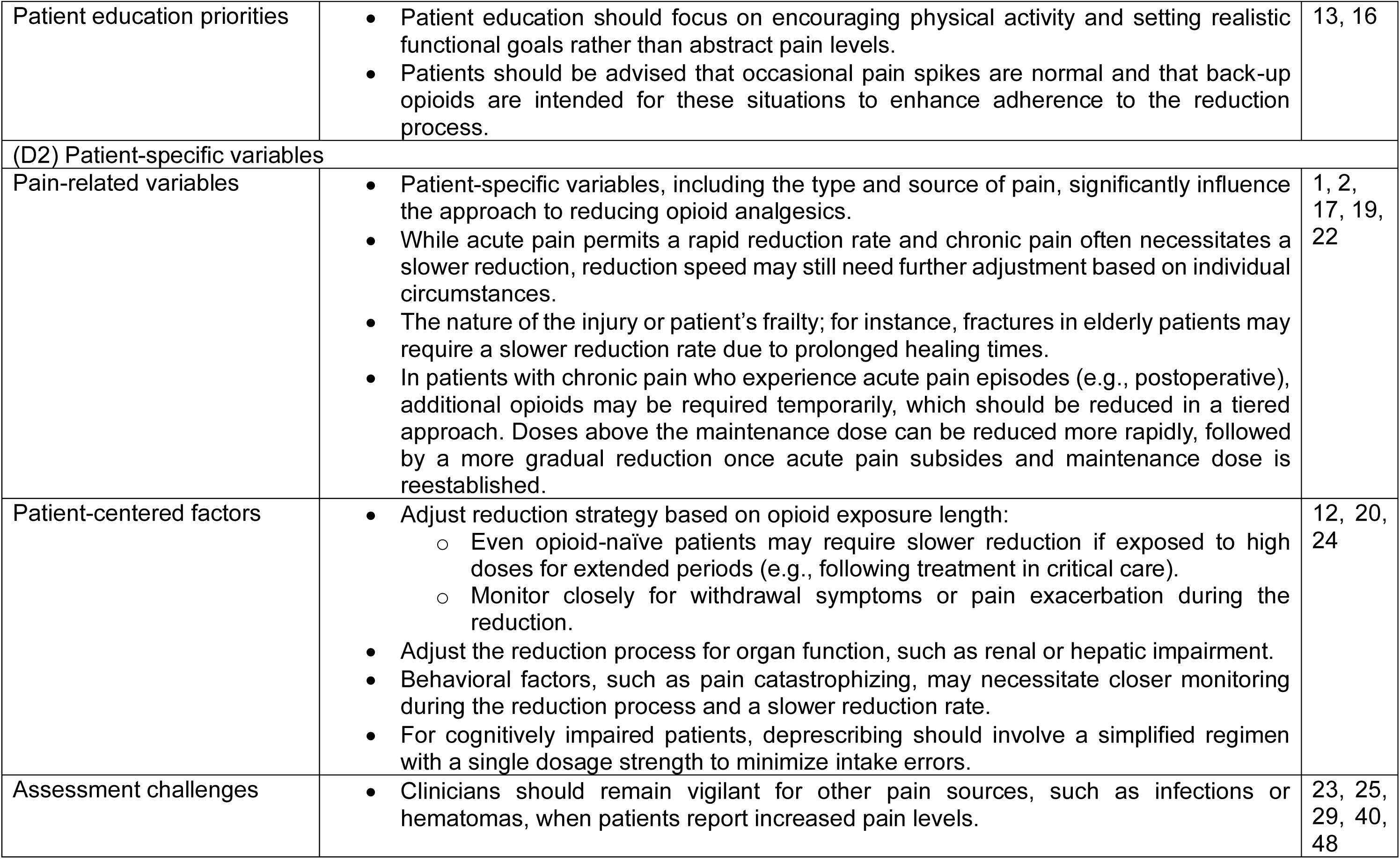
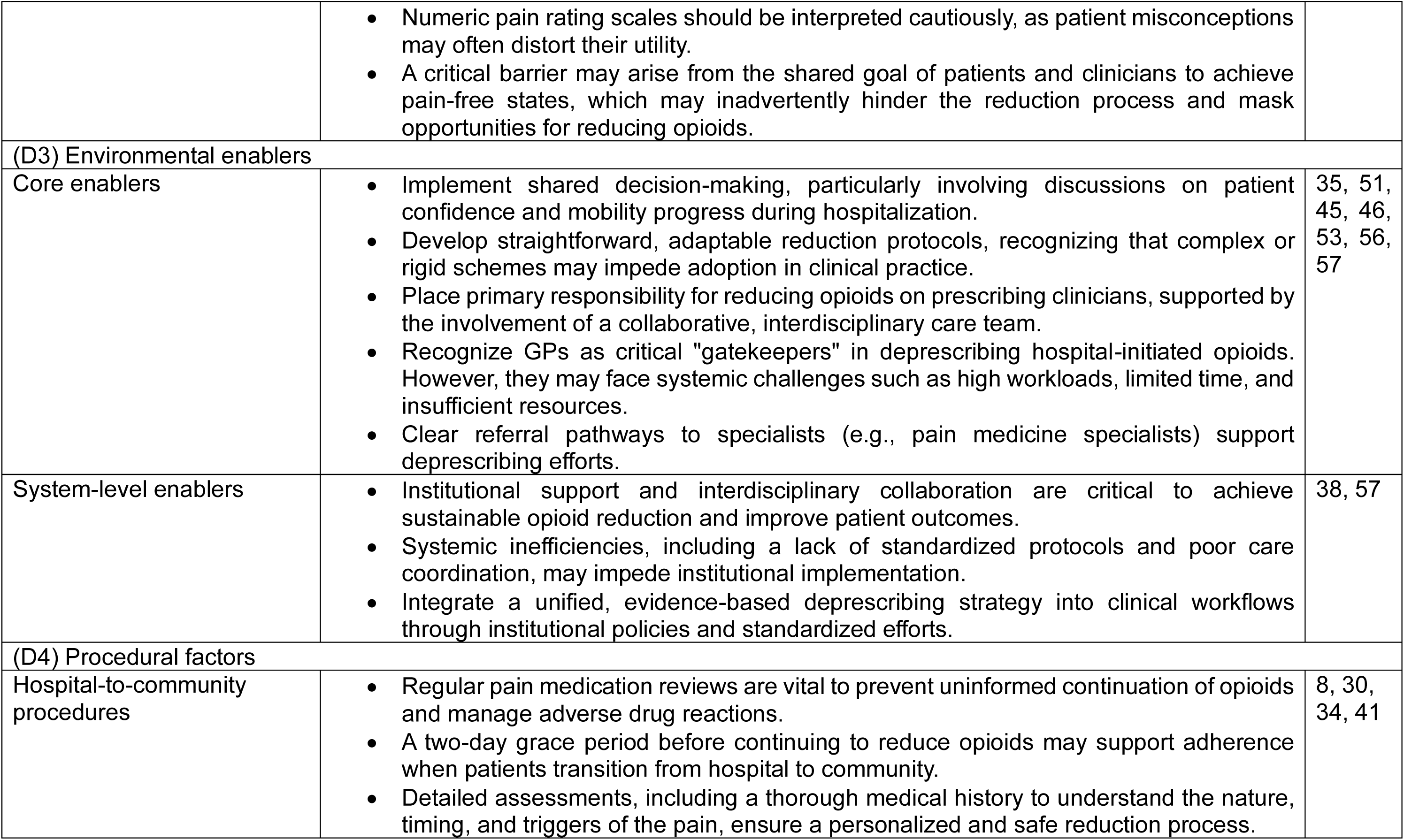
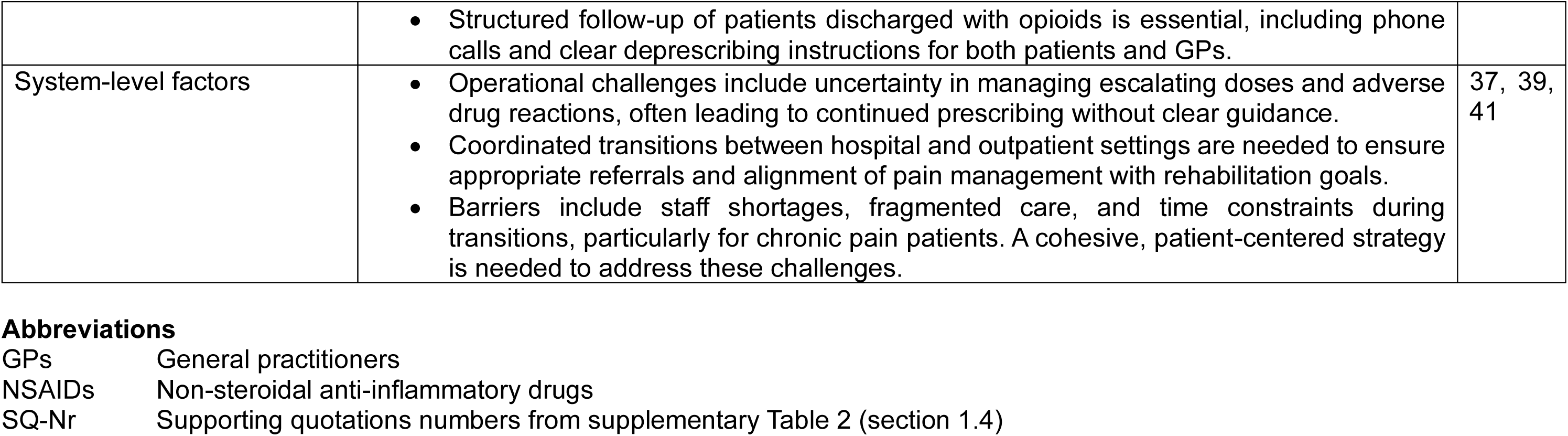
Four key domains described as needed for successful opioid dose reduction in hospitalized patients, derived through text condensation and examined through interaction analysis from focus group discussions and a Delphi survey conducted with local clinicians. For an improved overview, the respective domain contents are divided in subheadings. The corresponding supporting quotations are numbered and detailed in the supplement (section 1.4).

| Domain | Condensed recommendations | SQ-Nr |
| --- | --- | --- |
| (D1) Opioid reduction strategies |  |  |
| Structured reduction plan components | • Plan for opioid reduction whenever opioids are initiated or doses increased.<br>• Target either return to prehospitalization dose or complete cessation based on patient's prehospital opioid status (user vs. naïve).<br>• Commence reduction once patient stabilizes on regimen (minimal back-up opioid use and stable pain for 24-48 hours) and base starting deprescribing dose on patient's last 24-hour consumption of both fixed and back-up opioids.<br>• Adjust reduction rates (5-25%) based on pain type and nature of injury/surgical procedure to minimize withdrawal symptoms and maintain functionality:<br>○ Acute pain: 25% reduction with 2-day stabilization periods.<br>○ Chronic pain: 10% reduction with 3-day stabilization periods.<br>• For musculoskeletal injuries/surgeries, align reduction with mobilization process and use physical therapist input to guide reduction rates.<br>• Identify high-pain periods (e.g., morning activation) to prioritize which doses to reduce last, while maintaining back-up opioids at 1/10 to 1/6 of initial dose throughout reduction process.<br>• Reduce or stop opioids before discontinuing non-opioid analgesics (NSAIDs, acetaminophen, metamizole). There is reportedly a tendency to discontinue non-opioids first due to the higher pill burden required for adequate pain control in musculoskeletal conditions. | 1, 2, 5, 6, 7, 15, 19 |
| Barriers and solutions | • Large pack sizes and dosage forms of commercially available opioids, such as excessively high tablet counts or inconvenient administration routes, impede reduction efforts.<br>• Solutions include offering smaller, customized pack sizes and switching opioids to weaker agents or more manageable forms, such as tablets or capsules. | 10, 27, 28 |
| Patient education priorities | • Patient education should focus on encouraging physical activity and setting realistic functional goals rather than abstract pain levels.<br>• Patients should be advised that occasional pain spikes are normal and that back-up opioids are intended for these situations to enhance adherence to the reduction process. | 13, 16 |
| (D2) Patient-specific variables |  |  |
| Pain-related variables | • Patient-specific variables, including the type and source of pain, significantly influence the approach to reducing opioid analgesics.<br>• While acute pain permits a rapid reduction rate and chronic pain often necessitates a slower reduction, reduction speed may still need further adjustment based on individual circumstances.<br>• The nature of the injury or patient's frailty; for instance, fractures in elderly patients may require a slower reduction rate due to prolonged healing times.<br>• In patients with chronic pain who experience acute pain episodes (e.g., postoperative), additional opioids may be required temporarily, which should be reduced in a tiered approach. Doses above the maintenance dose can be reduced more rapidly, followed by a more gradual reduction once acute pain subsides and maintenance dose is reestablished. | 1, 2, 17, 19, 22 |
| Patient-centered factors | • Adjust reduction strategy based on opioid exposure length:<br>○ Even opioid-naïve patients may require slower reduction if exposed to high doses for extended periods (e.g., following treatment in critical care).<br>○ Monitor closely for withdrawal symptoms or pain exacerbation during the reduction.<br>• Adjust the reduction process for organ function, such as renal or hepatic impairment.<br>• Behavioral factors, such as pain catastrophizing, may necessitate closer monitoring during the reduction process and a slower reduction rate.<br>• For cognitively impaired patients, deprescribing should involve a simplified regimen with a single dosage strength to minimize intake errors. | 12, 20, 24 |
| Assessment challenges | • Clinicians should remain vigilant for other pain sources, such as infections or hematomas, when patients report increased pain levels. | 23, 25, 29, 40, 48 |
|  | • Numeric pain rating scales should be interpreted cautiously, as patient misconceptions may often distort their utility.<br>• A critical barrier may arise from the shared goal of patients and clinicians to achieve pain-free states, which may inadvertently hinder the reduction process and mask opportunities for reducing opioids. |  |
| (D3) Environmental enablers |  |  |
| Core enablers | • Implement shared decision-making, particularly involving discussions on patient confidence and mobility progress during hospitalization.<br>• Develop straightforward, adaptable reduction protocols, recognizing that complex or rigid schemes may impede adoption in clinical practice.<br>• Place primary responsibility for reducing opioids on prescribing clinicians, supported by the involvement of a collaborative, interdisciplinary care team.<br>• Recognize GPs as critical "gatekeepers" in deprescribing hospital-initiated opioids. However, they may face systemic challenges such as high workloads, limited time, and insufficient resources.<br>• Clear referral pathways to specialists (e.g., pain medicine specialists) support deprescribing efforts. | 35, 51, 45, 46, 53, 56, 57 |
| System-level enablers | • Institutional support and interdisciplinary collaboration are critical to achieve sustainable opioid reduction and improve patient outcomes.<br>• Systemic inefficiencies, including a lack of standardized protocols and poor care coordination, may impede institutional implementation.<br>• Integrate a unified, evidence-based deprescribing strategy into clinical workflows through institutional policies and standardized efforts. | 38, 57 |
| (D4) Procedural factors |  |  |
| Hospital-to-community procedures | • Regular pain medication reviews are vital to prevent uninformed continuation of opioids and manage adverse drug reactions.<br>• A two-day grace period before continuing to reduce opioids may support adherence when patients transition from hospital to community.<br>• Detailed assessments, including a thorough medical history to understand the nature, timing, and triggers of the pain, ensure a personalized and safe reduction process. | 8, 30, 34, 41 |
|  | • Structured follow-up of patients discharged with opioids is essential, including phone calls and clear deprescribing instructions for both patients and GPs. |  |
| System-level factors | • Operational challenges include uncertainty in managing escalating doses and adverse drug reactions, often leading to continued prescribing without clear guidance.<br>• Coordinated transitions between hospital and outpatient settings are needed to ensure appropriate referrals and alignment of pain management with rehabilitation goals.<br>• Barriers include staff shortages, fragmented care, and time constraints during transitions, particularly for chronic pain patients. A cohesive, patient-centered strategy is needed to address these challenges. | 37, 39, 41 |

**Abbreviations**
GPs General practitioners
NSAIDs Non-steroidal anti-inflammatory drugs
SQ-Nr Supporting quotations numbers from supplementary Table 2 (section 1.4)

Figure 2 denotes the identified key concepts of the determinants and their relationships. The implementation sequence begins with patient identification and engagement of an interprofessional healthcare team using standardized protocols, followed by establishing evidence-based reduction rates and target doses through collaborative clinical decision making. The reduction protocol is then customized according to patient-specific variables that may require further individualization of the approach. The individualized reduction strategy is subsequently communicated to both the case-leading physician and patient through structured educational materials. Finally, systematic follow-up is implemented to facilitate adherence to the reduction protocol and to monitor patient responses. A cross-section of thematic relationships, supporting participant quotations, and Delphi consensus items are included in the supplement (sections 1.3, 1.4, and 3.1).

**Figure 2.**
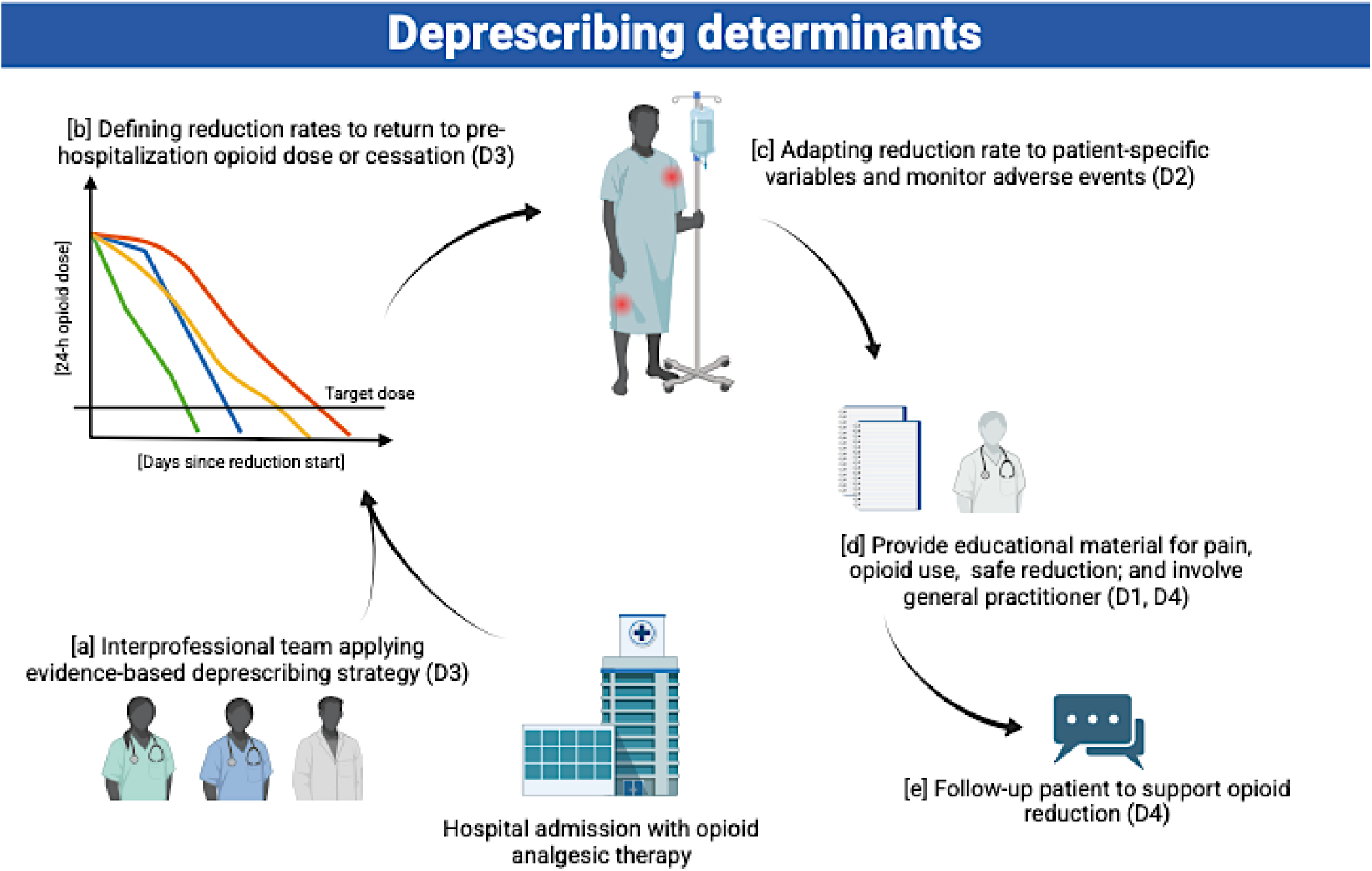
The figure illustrates the sequential implementation of identified opioid deprescribing determinants (D1 to D4) in the hospital setting. The numbering denotes the identified deprescribing determinants. The process begins with patient identification and [a] the engagement of an interprofessional healthcare team using standardized protocols. Next, [b] evidence-based reduction rates and target doses are established through collaborative clinical decision making. The reduction protocol is then [c] customized according to patient-specific variables that may require further individualization of the approach. The individualized reduction strategy is [d] communicated to both the patient and the case-leading physician through structured educational materials. Finally, [e] systematic follow-up procedures are implemented to facilitate adherence to the reduction protocol and to monitor patient responses.

### Opioid reduction calculator

As shown in Figure 3, the developed opioid reduction calculator proposes differentiated reduction rates based on opioid status: rapid rates for opioid-naïve patients and more gradual rates for those with chronic use. All approaches follow a linear reduction pattern to a patient’s specific target dose to enhance clinical adoption and predictability. The calculator’s interface enables the entry of opioid and back-up opioid use from the previous 24 hours as a starting point. For opioid rotation, it provides three reduction schemes that account for incomplete cross-tolerance, beginning with doses set at 30%, 50%, or 100% of the equianalgesic dose. This adaptability allows to select or switch between plans according to patient responses. The calculated doses can then be converted to commercially available formulations. The reduction plans can be further individualized based on different clinical parameters: organ function, surgery or injury severity, and functional goals, aiming to reduce opioids as consistently as possible to hospital preadmission levels. Supplementary Figure 2 (section 2.1) shows the calculator interface.

**Figure 3.**
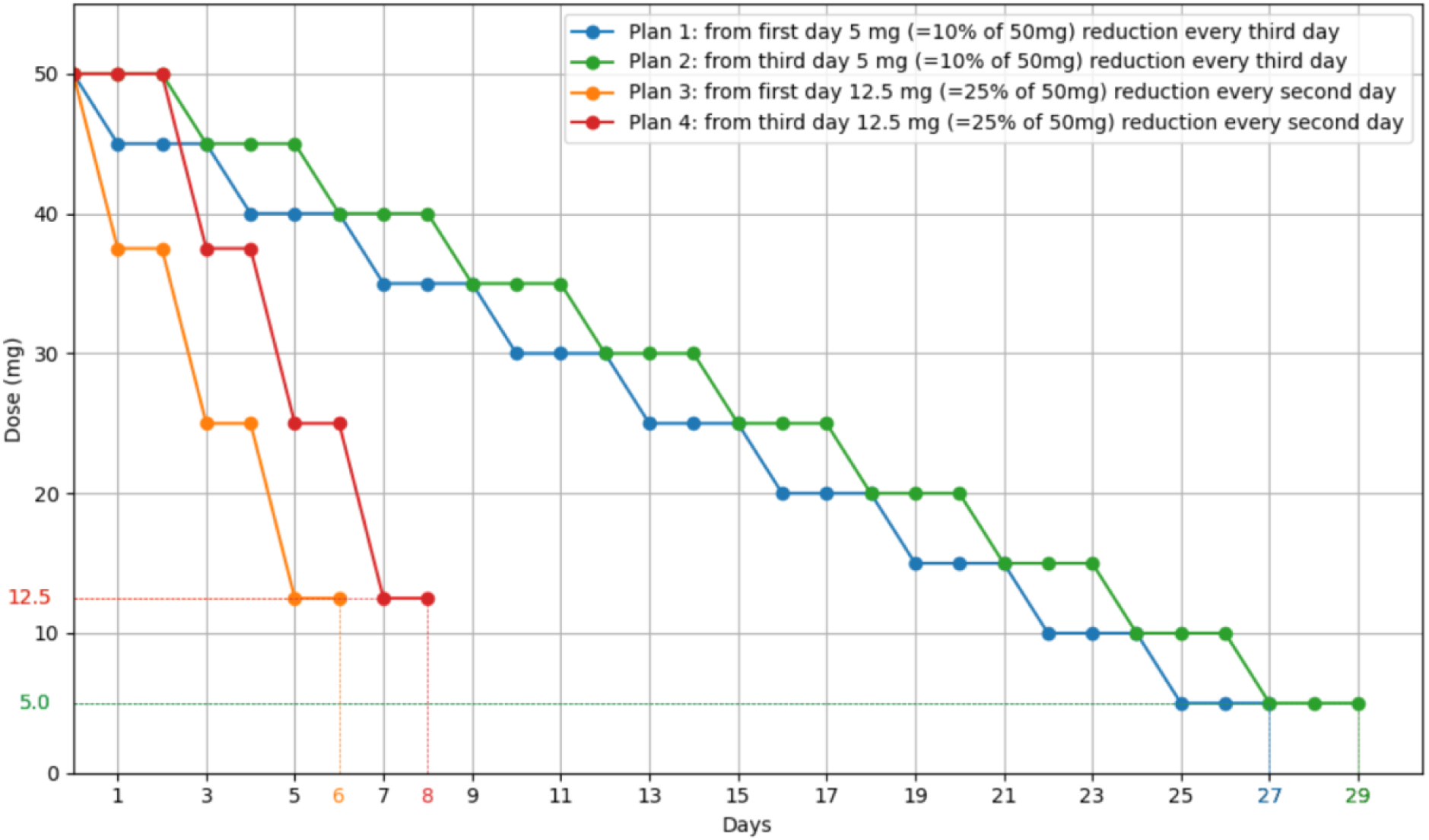
Derived from focus group discussions and a Delphi survey, four distinct reduction trajectories were delineated for reducing opioid analgesics in hospitalized patients, starting from a baseline dose of, e.g., 50 mg oral morphine equivalents and tailored to different clinical scenarios. For patients requiring a more gradual approach, plans 1 and 2 follow a 10% dose reduction every third day. For patients deemed suitable for rapid reductions, plans 3 and 4 reduce the dosage by 25% every other day, starting on the day one. Plans 2 and 4 also include a two-day grace period to account for opioid dose stabilization to accommodate reduction efforts if the patient is discharged on day 1. The calculated doses then need to be adjusted to commercially available formulations at the discretion of a healthcare professional.

### Patient interviews and information material

The opioid patient pamphlet included statements on opioid administration, therapy duration, alternative pain management, management of common adverse effects, and safe storage and disposal. We interviewed 13 patients to evaluate the pamphlet and 10 patients for the reduction handout (three were lost to follow up), with two participants being non-native German speakers. All patients correctly answered the comprehension questions. The mean time to read the pamphlet was 11.54 minutes (SD 5.16 minutes), and the mean comprehensibility score was 4.77 (SD 0.60). Patients suggested reducing subordinate clauses, keeping the text concise, using more conversational language, and providing clearer information about additional pain management activities. Some found details on regulations concerning travelling with opioids confusing and recommended deletion. Patients suggested simplifying explanations of adverse effects, adding more information on the duration of the analgesic effect, and formatting the pamphlet into a brochure size rather than the draft A4. Compared to the original sample, the number of sentences was reduced (-27, - 37%), as were the number of words (-377, -44%) and the number of syllables (-886, - 51%). All statements of the opioid patient pamphlet reached a Flesch Reading Ease Score of 43, which is indicative of an average newspaper (43). For the reduction plan, the comprehensibility score was 4.50 (SD 0.85), with some requiring multiple readings. Patients suggested focusing more on opioid analgesics rather than other pain medications used to reduce opioid doses, using fixed times instead of intervals such as “every 8 hours”, and providing guidance on whether opioids should be taken with or without food. All patients stated they would recommend the pamphlet and the reduction plan to other patients.

## Discussion

This multi-level consensus study identified four key components of a successful opioid deprescribing framework for a systematic reduction of opioids in tertiary care: (1) deprescribing determinants consisting of actionable reduction rates that are implementable in a reduction calculator; (2) patient-specific variables to be considered for successful opioid deprescribing; (3) contextual factors critical for a locally accepted implementation of a standardized opioid reduction strategy; (4) patient-centered educational content on opioids and dose reduction. The initial focus group findings identified broad applicability across medical specialties, as confirmed through the achieved cross-specialty consensus in a Delphi survey spanning six medical disciplines. We translated these findings into a calculator that proposes individualized reduction plans for various opioids and different clinical parameters. Importantly, patients reported that the educational elements were both comprehensible and valuable in supporting their opioid reduction process.

Patients receiving new or escalated opioid doses face considerable risks of transitioning to long-term use and experiencing poorer pain outcomes (4). While evidence for identifying suitable candidates for opioid reduction has expanded, current prescribing and deprescribing guidelines (15–18) universally recommend that a reduction plan should be laid out for all patients with escalated doses. While these guidelines provide structured (17,18) or conceptual guidance (15,16), these recommendations lack end-to-end considerations of integration into existing healthcare settings from clinician and patient perspectives. This gap between recommendation and practical application potentially contributes to clinicians’ lack of self-efficacy in managing opioid reduction. Previous studies examining multidisciplinary hospital clinicians through focus group interviews (44,45) revealed challenges in incorporating deprescribing into medication reviews, specifically noting the absence of tools to guide confident reduction trajectories. Our findings align with this identified need, with participants endorsing the value of a calculator proposing an initial reduction plan. Rather than filling this gap solely with automated clinical decision support, our study suggests that successful prevention of long-term opioid dependence requires a dual approach: patient-centered reduction strategies supported by robust institutional frameworks.

As the systematic review of OEPs had identified (8), the participants in our consensus approach described effective reduction of opioids to be largely dependent on tailoring the approach to each patient’s clinical characteristics. These included the type of pain, duration of opioid exposure, functional goals, age, and overall health status. Opioids used for acute pain, such as after surgery or trauma, should allow for rapid reduction rates up to 25%, whereas chronic pain requires slower reduction rates of 5 to 10% to avoid withdrawal symptoms and maintain physical activity. This finding is consistent with previous findings that there are different withdrawal phenotypes that require different reduction strategies due to different withdrawal symptom presentations (46). Organ function, including hepatic and renal health, must also guide reduction rates, as impaired metabolism or excretion of opioids or their active metabolites can prolong half-lives (47,48). As pain is recognized as a biopsychosocial response (49), behavioral aspects such as pain catastrophizing can inform the need for individualized reduction strategies to select slower reduction rates and support adherence to the plan. Similarly, psychological comorbidities were identified as essential for planning pain management and opioid prescribing (15,16,18). Encouraging patients to focus on functional recovery rather than achieving pain-free states is critical, as functional goals provide a tangible goal for progress and increase patient engagement in the process. These efforts can be complemented by follow-up calls and educational materials to guide patients through the reduction process.

In addition to patient characteristics, participants agreed that a supportive environment and systematic engagement are critical to successfully reduce opioid use. Structured follow-up care with regular medication reviews emerged as essential for tracking progress and addressing challenges during the reduction process. The transition from hospital to community care proves particularly critical, requiring clear, actionable deprescribing instructions for both patients and GPs. However, systematic barriers, including fragmented care, staff shortages, and high workloads, may undermine these efforts. Institutional support, such as standardized protocols and accessible specialist consultations, can address these gaps and facilitate a coordinated approach. Langford and colleagues previously described similar barriers as health system-related challenges of opioid deprescribing (45). As a solution, our participants emphasized the need for interdisciplinary collaboration, particularly during hospitalization, where physiotherapists and pain specialists can provide critical input on mobilization targets and pain management goals. Additionally, engaging patients through shared decision-making can foster trust and adherence by aligning reduction plans with their individual circumstances and expectations. These findings highlight the importance of a cohesive, patient-centered strategy that integrates evidence-based guidelines into routine practice, reinforcing the broader role of opioid stewardship initiatives (7) in reducing long-term opioid use. We addressed these issues by developing a deprescribing framework, reduction calculator, and handouts to follow, which were recently identified as enablers of opioid deprescribing in interviews with GPs (50).

This study has important limitations. Participants for the interviews and the Delphi survey were recruited from a single institution, which introduces a potential sampling bias. Only one focus group discussion was conducted, which did not allow a comprehensive assessment of data saturation. As a result, it is possible that some issues or perspectives were incompletely captured. However, this study represents an attempt to adapt evidence and international guidelines to local conditions and the experience of the prescribing medical specialties. This means that the findings may not be directly applicable to other institutions, limiting their generalizability. However, this study can guide other researchers and clinicians in developing their own frameworks.

This study has notable strengths as well. It fostered, and showcases, open institutional discussions guided by the latest evidence and tailored to local needs. The methodology incorporated a wide range of medical expertise, blending insights from both senior and early-career physicians. The robustness of our focus group findings was substantiated through subsequent validation through a Delphi survey, which exposed the findings to a broader range of professionals that may later be affected by its implementation. We reached full consensus and stability across the synthesized recommendations. Patient involvement further allowed us to refine the strategies used to communicate a reduction of opioid analgesics, establishing a holistic framework for opioid deprescribing in tertiary care.

### Future outlook for clinical practice

Our findings help move theoretical recommendations to practical implementation and institutional-specific opioid deprescribing protocols in tertiary care settings. The identified environmental and procedural factors can guide the implementation process to meet the requirements necessary for integration into clinical practice, whilst the identified patient-centered determinants demonstrated broad applicability, requiring minimal modification for widespread adoption. Figure 4 shows a practical blueprint for a collaborative workflow between an opioid deprescribing service and the case-leading physician, derived from our established opioid deprescribing framework. Such a workflow may facilitate systematic opioid reduction while maintaining continuity of care from hospital admission through the hospital-to-community transition.

**Figure 4.**
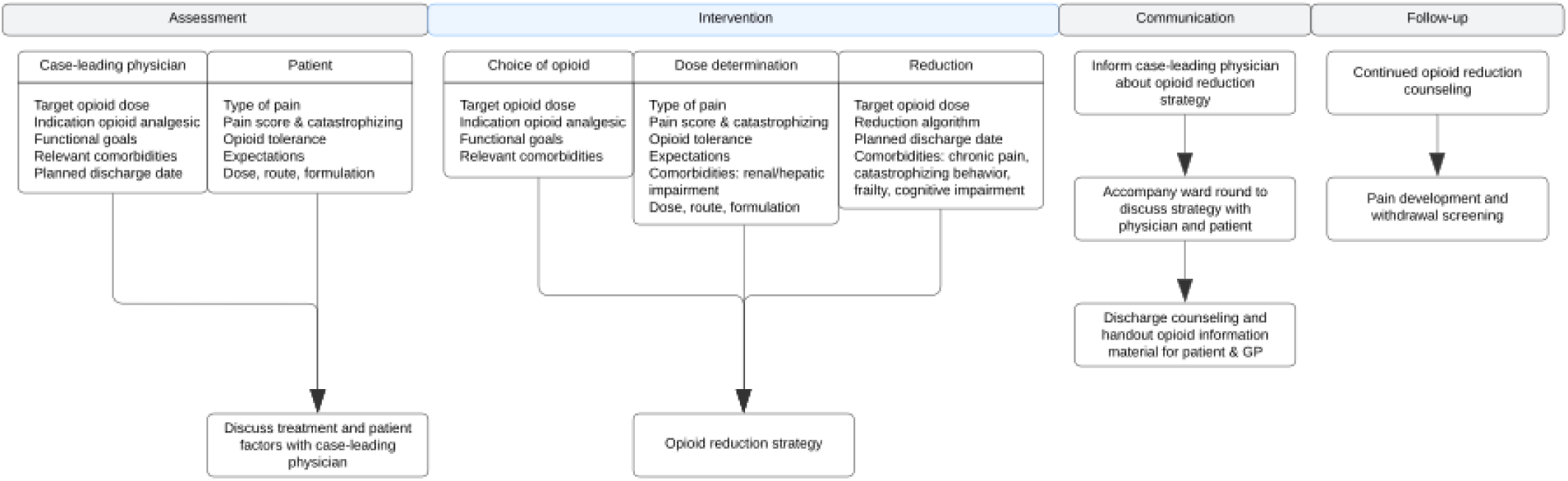
A practical blueprint for a collaborative workflow between an opioid deprescribing service and the case-leading physician to reduce opioid analgesics in hospitalized patients with planned reduction or end of opioid therapy. GP = General practitioner.

## Conclusions

Through the systematic integration of evidence-based guidelines, clinical expertise, and patient perspectives, we developed a local opioid deprescribing framework that bridges the gap between theoretical recommendations and practical implementation. The framework’s adaptability to diverse clinical scenarios, combined with its patient-centered approach to opioid dose reduction, provides healthcare institutions with an actionable blueprint for opioid stewardship. Our multi-level consensus approach demonstrates the value of collaborative engagement in developing clinically viable protocols for complex medication management. The next step involves conducting prospective, confirmatory trials to determine the effect of adopting this framework on opioid analgesic use and patient-reported outcomes relevant for medication withdrawal.

## Availability of data and materials

All data generated or analyzed during this study are included in this published article and its supplementary materials, except for the interview transcripts and Delphi survey data. Due to the potential risk of participant identification, access to the transcripts and Delphi survey data will be provided only upon reasonable request to protect their anonymity.

## List of abbreviations

CFIR: Consolidated framework for implementation research CI Confidence interval
COREQ: Consolidated criteria for reporting qualitative research GP General practitioner
OEP: Opioid exit plan

## Acknowledgements

We thank all interview and survey participants for their time and for providing informative and helpful responses. We also thank the Department of Pain Medicine at the University Hospital Basel, whose patient pamphlet we could use as material for our study. We used Deepl Translator (DeepL SE, Köln, Germany, version 24.11.21416769) for translation of specific text passages from German to English. GPT-4o mini was used during the preparation of this work to improve readability. After using these tools, the authors reviewed and edited the content as needed and take full responsibility for the content of the publication.

## Funding

This study was supported by an external research grant “Forschungsprojekt nationaler Tragweite 2023” from the Swiss Association of Public Health Administration and Hospital Pharmacists (GSASA). Open access funding was provided by ETH Zurich.

## CRediT author contributions

MR: Conceptualization, methodology, validation, formal analysis, investigation, data curation, writing – original draft, visualization, project administration, funding acquisition. ES: Software, validation, formal analysis, investigation, visualization, writing – review & editing. MMM: Conceptualization, project administration, writing – review & editing, funding acquisition. AMB: Conceptualization, writing – review & editing, supervision, funding acquisition. DS: Conceptualization, methodology, formal analysis, investigation, writing – review & editing, supervision, funding acquisition.

## Ethics declarations

### Ethics approval and consent to participate

Ethical approvals were sought from ETH Zurich Ethics Commission (EK-2024-N-37, Project 24 ETHICS-270). All study participants provided written informed consent.

## Competing interests

The authors declare that they have no competing interests.

## Notes

### Competing Interest Statement

The authors have declared no competing interest.

### Author Declarations

Institutional ethical approval was obtained from the ETH Zurich Ethics Commission (EK-2024-N-37; Project 24 ETHICS-270). All research was performed in accordance with the Declaration of Helsinki. Written informed consent was obtained from all study participants.

### Summary of Updates

In the initial submission to medRxiv, the supplementary material mentioned in the manuscript was inadvertently omitted. The submission has now been revised to include the supplementary material. In addition, high-resolution figures are now available. The manuscript itself has not been altered.

