## Supplement for "Optimizing hospital opioid deprescribing: a multi-level consensus study bridging evidence, expertise, and patient perspectives"

### 1. Focus group details

The focus group interview was conducted by a research team comprising one male pharmacist and senior researcher (DS: facilitator and co-interviewer, MSc, PhD), one male pharmacist and PhD student (MR: interviewer, MSc), and one female pharmacy master's student (ES: non-verbal note-taker, BSc). All members of the research team and participants worked at the same hospital and were therefore acquainted. Only the participants and research team were present during the discussion. The facilitator and interviewer disclosed their professional interests but refrained from sharing personal assumptions to minimize bias. The interview guide was pilot-tested before use.

Participants were briefed beforehand about the purpose and goals of the focus group. Recruitment was conducted via telephone or email. When one participant declined due to a scheduling conflict, a substitute was invited to ensure full participation. No repeat interviews were conducted. Instead of relying on data saturation, the findings were further refined and validated through a Delphi survey involving a broader group of medical professionals.

#### 1.1 Focus group interview guide

**Tasks/Responsibilities:**

<u>MR:</u> Room reservation, food/refreshments organization, moderator
<u>ES:</u> Technician (audio recording), note-taker (seating order, non-verbal expressions)
<u>DS:</u> Timekeeper, co-moderator

**Settings:** Bright room with large screen, tables ordered in a U-shape

**Materials:**

- Audio recording (plus backup recorder)
- Timer for sections
- Projector screen for PowerPoint presentation
- White board with differently colored pens
- Pens and paper for the participants
- Water bottles for the participants

**Time table:**

| | |
|---|---|
| 15:30 - 15:35 | Arrival and provision of food/refreshments |
| 15:35 - 15:45 | Participant and researcher introductions |
| 15:45 - 15:55 | Topic introduction |
| 15.55 - 17:30 | Focus group discussion |
| 17:30 - 18:00 | Apéro |

**Interview questions have been translated from German to English**

*Introductory questions*

1. How do you experience the use of opioid analgesics in your everyday life?
2. Case study: Female patient with osteoarthritis, elective total hip replacement, arterial hypertension, possible CHD, TEE 03.2022, and moderate depressive episodes, treated with escitalopram 10 mg

*Probing questions*

3. Case study discussion: how would you manage the pain and continue analgesic therapy?

*Explorative questions*

4. When is the ideal time for you to start tapering with a patient?
5. How quickly should an opioid be reduced?
6. Who is responsible for implementing the tapering model?
7. Can a reduction scheme support the transition from hospital to home?
8. Review of three published opioid reduction plans
    a. Tamboli et al., 2020; Joo et al., 2020; Kukushliev et al., 2023 [1–3]
    b. Genord et al., 2017 [4]
    c. Chen et al., 2020 [5]
9. What do you like about these templates?
10. What do you dislike about these templates?

*Closing questions*

11. Which 3 to 5 key elements should a tapering tool contain?
12. Which types of pain should the reduction scheme be used for?
13. What can support the rehabilitation process during the opioid reduction phase?

Structure of the focus group discussion

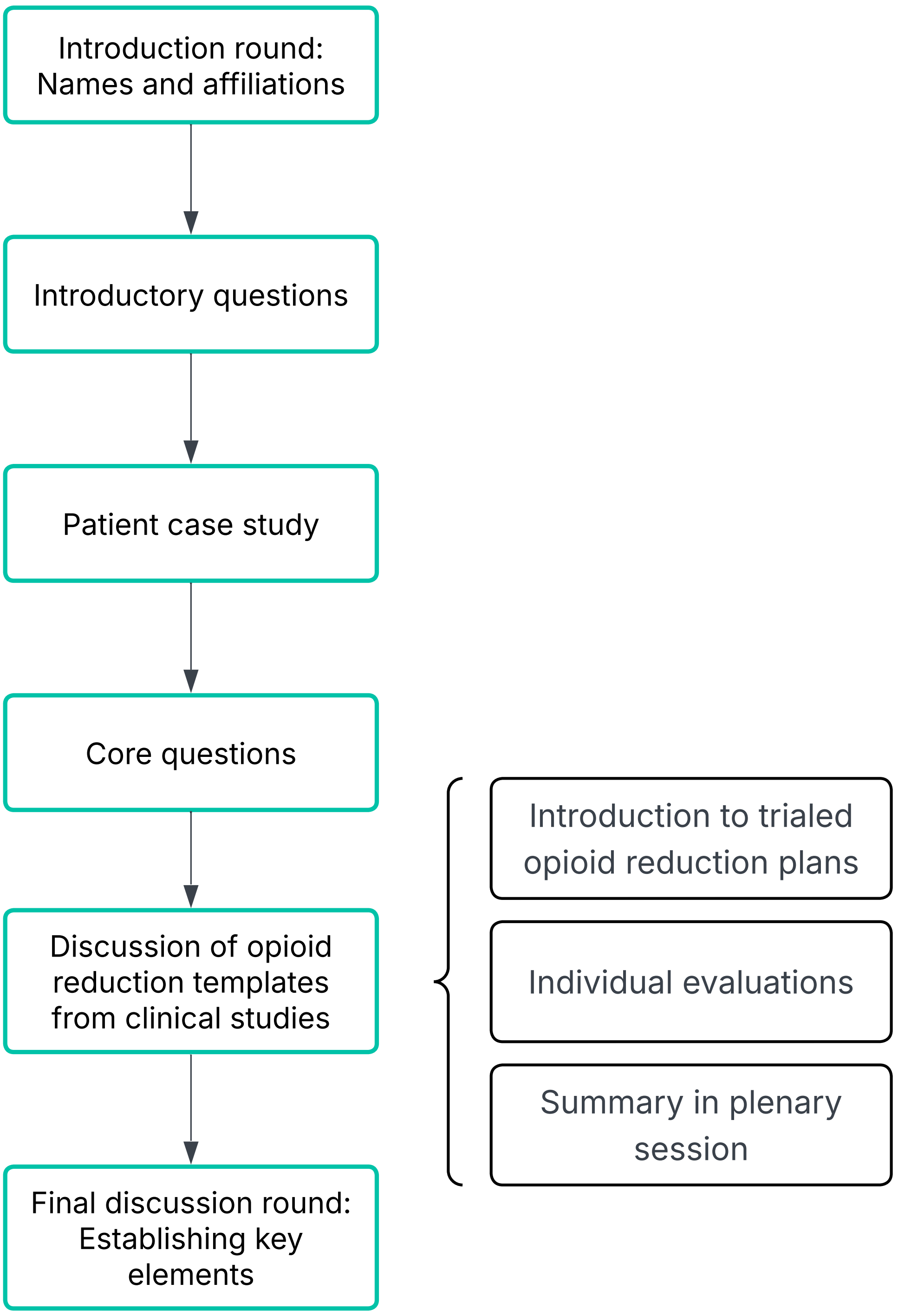

#### 1.2 Focus group content analysis: coding tree

**Table 1** The coding tree for deductive content analysis was clustered into four domains, five major themes, and 15 subthemes. A definition for uniform coding is provided in the last column.

| Domain | Theme | Subtheme | Code definition |
|---|---|---|---|
| **Intervention characteristics:** Describing the intervention characteristics involves detailing how the tapering plan/protocol should appear, identifying its key elements, and outlining what the tapering process should cover. | Tapering plan | Reduction rate | When tapering is indicated, consideration should be given to the rate at which the patient reduces their opioid dosage, with the aim of avoiding withdrawal symptoms. Also, it includes at which dosage abrupt cessation is acceptable. |
| | | Duration | Regarding the tapering process, how long should it take to achieve the desired opioid dose, which could be zero or another specified goal? |
| | | Initiation of tapering | What parameters suggest that it's appropriate to initiate the tapering process? When is an appropriate time to begin tapering? |
| | | Adaptability | How should the tapering plan/protocol accommodate diverse patient histories? How can it adjust to variable factors? How can and should the tapering process be adapted to changing circumstances, such as the appearance of withdrawal symptoms? |

| | | | |
|---|---|---|---|
| | | Design | How should the tapering plan/protocol be structured, and what elements should be included? Should it be in electronic or paper form?<br>It should also include which active pharmaceutical ingredients the tool can be used with. |
| | | Recipient | Who is the intended audience for the tapering plan, and who needs to comprehend it? Should both physicians and patients receive identical plans? |
| | Patient pain education | Patient pain education in paper form | What additional materials, apart from the tapering plan/protocol, should the patient receive to be informed about their opioid use/opioid tapering? |
| | | Patient pain education in person | What topics should be discussed with the patient at hospital discharge, and how should this information be communicated? What are the key aspects of the patient-physician interaction? |
| **Patient characteristics:**<br>It involves patient characteristics, which can influence opioid prescription and tapering. This includes identifying factors that may require a tailored tapering approach. | Individualized patient care | Type of pain | How can the type of pain (e.g., neuropathic vs. nociceptive, or acute vs. chronic) affect the tapering process? |
| | | Opioid exposure | How should the tapering process differ between an opioid-naive patient and an opioid-exposed patient? |
| | | Other clinical parameters | Do other clinical parameters, such as comorbidities, influence the tapering process? |
| | | Barriers | What patient factors contribute to the difficulty of the opioid prescription/tapering process? |

| | | | |
|---|---|---|---|
| **Environment:** The environment involves the context in which the tapering tool is situated. This includes the current state of pain management at the cantonal hospital in Baden, as well as the broader external setting such as general practice offices. It also addresses future considerations, such as desired outcomes and the necessity for opioid tapering assistance. | Hospital pain management | Postoperative-Status-quo | How is postoperative pain management conducted in tertiary care currently? |
| | | Postoperative-Future | Desired improvements in postoperative care, which could enhance patient care, which are presently unpracticed but are hoped for. |
| | | Standard of care-Status-quo | What general factors are considered in tertiary care pain management currently? What influences the prescription of pain medications, especially opioids currently? It includes both pharmacological and non-pharmacological interventions. |
| | | Standard of care-Future | What factors in tertiary care pain management are currently not practiced but would be beneficial if implemented in the future? How should the standard of care of pain management, with the focus on opioids should look like in the future? What are the expectations for the future? It includes both pharmacological and non-pharmacological interventions. |
| | | Barriers | What challenges could we face with implementing a tapering tool in tertiary care? |
| | | Areas for improvement | What is currently lacking regarding opioid therapy in tertiary care, and do the physicians perceive a need for tapering assistance? |

| | | | |
|---|---|---|---|
| | External setting | General practitioner-Statusquo | What is the general practitioner currently occupied with? Describe their current situation and what they are currently doing. It involves how general practitioners are perceived and what their abilities are. It also includes the problems currently arising at the general practitioner. |
| | | General practitioner-Future | What is expected from a general practitioner? |
| **Process:** The process involves how the tapering process should work, what the stages will be, and who will participate. | Process | Clinician responsibility | Who carries the responsibility of the tapering process? |
| | | Communication | Considerations regarding communication among different healthcare professionals and the transition between various departments (e.g., between hospitals and general practitioners, or within the hospital). |
| | | Barriers | What are the current challenges facing the tapering process, and what difficulties could arise with the tapering protocol? What are the challenges for implementing the tapering tool? |
| | | Interdisciplinarity | Who should be involved in the tapering process, and what tasks should each participant perform? |

| |
|---|
| **Category X:** This is the code for all segments that are not relevant to our research question or are random statements. |

#### 1.3 Thematic relationships

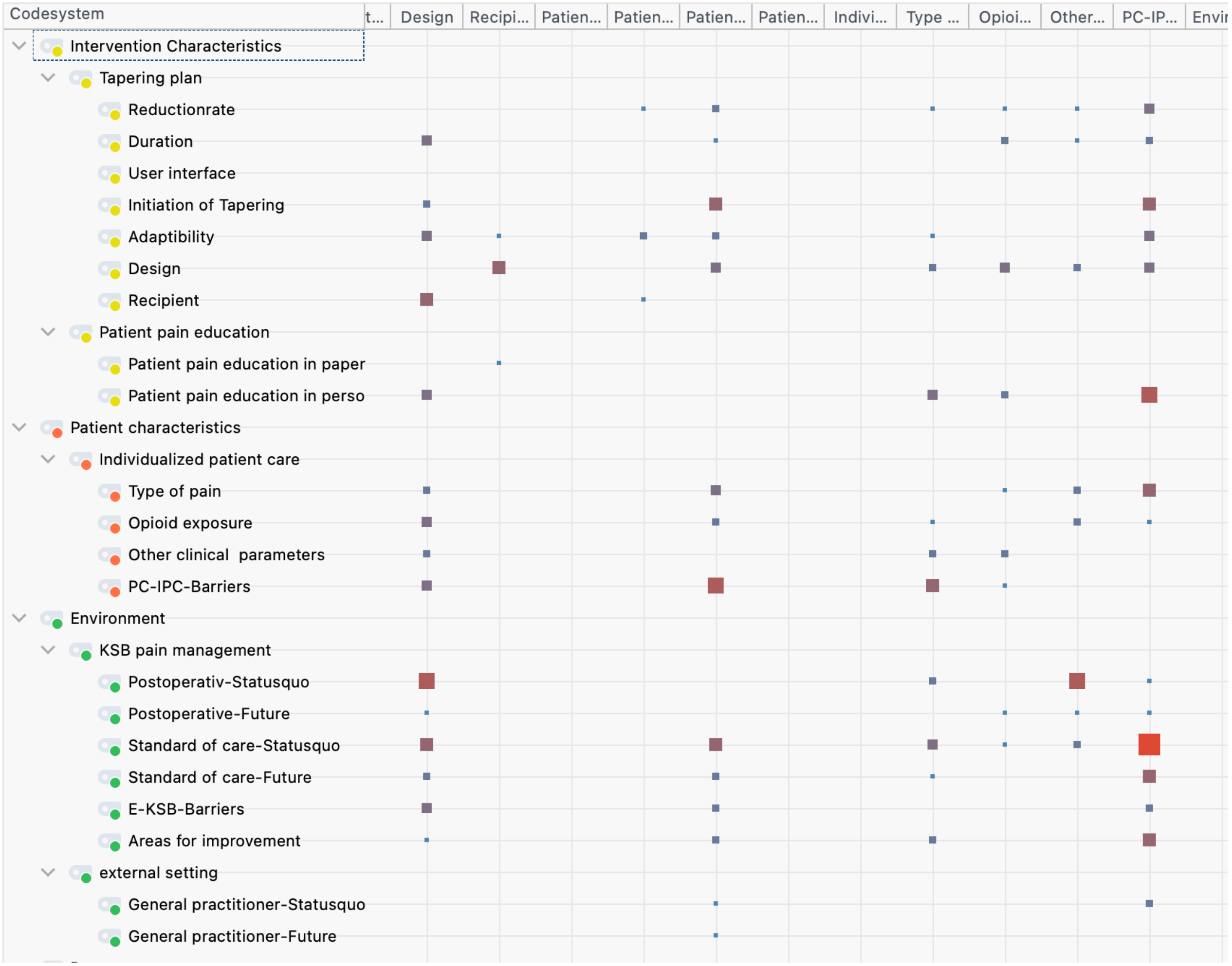

**Figure 1** The strength of the relationship is shown as a result of the interaction analysis. Strength was defined as the distance between the codes in the transcript.

#### 1.4 Supporting quotations from the clinician focus group discussion

**Table 2** Supporting quotations from the clinician focus group discussion used to construct the deprescribing determinants and opioid reduction calculator are indicated by their corresponding coded subtheme.

| Nr | Subtheme | Interviewee quotations |
|---|---|---|
| 1 | Reduction rate | “Around five to 25%, which is probably the range, but it's quite individual. And there are also patients who stop taking everything overnight, which isn't great either.” |
| 2 | Reduction rate | “But they should already follow a certain pattern and try to go down 10% every three days, for example” |
| 3 | Duration | “On the other hand, why? 10 days is not yet chronic and not forever. So if they weaned off in 10 days, I'd be happy too. The main thing is that he weans off. I'm very pragmatic about it.” |
| 4 | Duration | “And that's why I would think that you (orthopedics) at least have this safety check with the six weeks and we don't, of course. But in the case of non-specific acute back pain, the six weeks would also be a time when you should actually look further if it hasn't gone away.” |
| 5 | Initiation of Tapering | “But I don't do it just like that, without asking the clinical question: How are you? I pay less attention to the pain scale and also what the curve says. How relaxed is he lying in bed? Was I perhaps in the room while the physiotherapist was mobilizing the patient and saw out of the corner of my eye how he was doing? How is he doing? Is his face contorted in pain or is he walking around in the corridor in between?” |
| 6 | Initiation of Tapering | “I think the (hospital) discharge depends on the mobilization in the end. So if they can't be mobilized, it can't be discharged. In other words, we can only start with the discharge once mobilization has taken place on our (orthopedics) part.” |
| 7 | Adaptability | “Is it sometimes worth asking when the pain is worse, during the day or at night? Maybe you can also start to reduce an evening dose when people are actually sleeping well, but when there's more movement, sometimes you do that a bit, just take one a day.” |
| 8 | Adaptability | “Give them 48 to 72 hours to establish themselves in the home setting, because they actually do everything themselves. They no longer have food delivered, they have to walk around themselves, they certainly move around more than in the hospital. And that they can check where they currently find themselves.” |
| 9 | Design | "Basically, the first page gives the result of the calculation in the background, what the patient has to do. If you had a tool, okay, like a drop-down menu. I have hydromorphone, I have what the vulture, so and so has the procedure. If you click, click, click and at the end it calculates it for you, then I think |

| | | |
|---|---|---|
| | | I've already saved time. But the work is done in the background and I don't have to do it myself with a spreadsheet. I don't think it would be bad if you could do it that way." |
| 10 | Design | "And that's why it should actually be something where you can reduce gradually with one medication. Yes, not one pack every time or if I take it for three days, 10, then three days, five, then I already have two packs of 30 tablets at home." |
| 11 | Recipient | "Maybe like this: Specify the baseline or the target value. Exactly. The target value, whether you really want to go to zero or whether you actually say that the previous dose was this and that, and that is perhaps our primary goal now." |
| 12 | Recipient | "I think a tapering scheme like this simply has to be as simple and understandable as possible for the patient, but also for us. Because we also need the time to explain it. And if it gets any more complicated, then it's difficult." |
| 13 | Patient pain education in paper form | "Shouldn't a reduction scheme also have something educational in it, such as non-pharmacological measures and (...) goals? Functional goals?" |
| 14 | Patient pain education in paper form | "A separate booklet "Dealing with pain" or something like that. Something simple. A leaflet." |
| 15 | Patient pain education in person | "I think we can often (draw attention) and I think it's important to communicate that the Dafalgan, the Novalgin, should not be stopped first, for example. Instead, the opiate should be stopped first and only then the others." |
| 16 | Patient pain education in person | "And that's why I want to take good care of the pick-up so that they don't have the problem. Okay, if they take it away from me now and I walk up the stairs, then it hurts so badly again, then I'm blocked, so I'm certainly not going to put it down. That brings us back to communication, that you also pick them up." |
| 17 | Type of pain | "And that's why acute pain patients are often not such a big issue after the operation, simply because they have a clear trigger. Or they have a scar, a prosthesis. But even there the willingness to reduce and stop is already very high. Unless they become chronic pain patients." |
| 18 | Type of pain | "That you also set achievable goals, because freedom from pain is unrealistic for chronic patients and I think you also have to set goals for acute patients, depending on their initial condition." |
| 19 | Opioid exposure | "I think there are two patients again, the opioid-naïve or the (opioid) chronic. The chronic one would be my personal goal now. He needs to at least get back to where he was before the intervention. That would be nice. Less would always be better because it might hurt less after the procedure. And |

| | | |
|---|---|---|
| | | the opioid-naïve one might be a bit quicker, because he's not as chronic yet and the opiate receptors aren't as triggered.” |
| 20 | Opioid exposure | “The short-stay patient, who needs less time than someone who is now in hospital for eight weeks in an intensive care unit, certainly needs more time, and I think he can have more time so that he doesn't become “drained”.” |
| 21 | Other clinical parameters | “I'm more concerned about where he's going. Is he going to physical rehab?” |
| 22 | Other clinical parameters | “Pelvic compression fractures take time and that hurts. Especially the older patients, they can't compensate so well, they might not have enough muscle mass or strength.” |
| 23 | Other clinical parameters | “What is still common is when it has bled (postoperatively).” |
| 24 | Other clinical parameters | “I really believe that how individuals deal with pain is crucial and that, if we go into a study and we will do so, we should also map this “catastrophizing” component and other avoidance behaviors. Because, firstly, they can be influenced and, secondly, they probably also play a role in this topic. I don't know to what extent we can implement this in a tapering scheme, but we can at least map it somehow.” |
| 25 | Barriers | “And that's the most difficult thing, because the patient naturally wants to be pain-free.” |
| 26 | Barriers | “I actually have the impression, I have to say, that people don't want to take tablets in the first place. They are happy (if they dont have to take them)... So that's also a key issue. With back pain, for example, we always say: “Stay active, go back to work, exercise and so on.” But people prefer not to take pills and lie in bed.” |
| 27 | Barriers | “Another problem with outpatients is that they suddenly need a different pack size, which they still have at home. Or maybe the fives. Or they have the 10s, then they have fives. And then you also have to make sure that they don't suddenly mix them up. And then there are 15 instead of 10 or five” |
| 28 | Barriers | “And that has something to do with the quantity of tablets. Or if you prescribe oxycodone/naloxone, for example, you have one in the morning and one in the evening. But metamizol, you have to take two, two, two. That's a lot of tablets and people naturally want to take fewer tablets in the first place.” |
| 29 | Barriers | “I think another important aspect is what you've just mentioned. Patients simply don't understand the abstract form of this pain scale (NRS).” |

| | | |
|---|---|---|
| 30 | Postoperativ-Statusquo | “I would first ask: “What is the pain like? Where is the pain? When does the pain happen?” All the 'W' questions and not just give a blanket answer. So you need a bit of an anamnesis so that you don't just give something like that, i.e. basic analgesia for sure, but you need an anamnesis. What, when, how, where does it hurt?” |
| 31 | Postoperative-Statusquo | “Exactly. And now he still has (an NRS) five to six. And of course you then check during the inpatient stay whether that's enough. So I or we don't like to prescribe opioids. I don't really let anyone go home with opioids unless they really have to. It's a bit different in the spine, but less so in orthopedics. And there you could still discuss whether you might want to prescribe Tramadol as a weak opioid. Yes.” |
| 32 | Postoperative-Statusquo | “So we don't really call patients and ask them: “And have you stopped taking opioids yet?”" |
| 33 | Postoperative-Future | “And I think it's also the case that it probably makes little sense to intervene at all in such a short episode. But it still sometimes leaves you with a guilty conscience when you think: “Yes, well, we should have tackled that now, shouldn't we?” Patients frequently come for something else.” |
| 34 | Postoperative-Future | “But it probably wouldn't be a bad idea to follow up with a phone call. I mean, there aren't that many patients who actually go home with opioids, so you could ask about it. So if you somehow (...) standardize that.” |
| 35 | Standard of care-Statusquo | “So it's not just a matter of deciding on your own, but in agreement with the patient, of course. How do they feel? How was the mobilization? What do they feel confident about, what don't they feel confident about? And, of course, what did he need during his stay in hospital?” |
| 36 | Standard of care-Statusquo | “Physical rehab. Then I might, I think to myself, hand out reserve opioids because I know that the patient is still under control. And I also want the rehab to make sense. I mean (an NRS of) six to seven, five to six, maybe it's gone down a bit on discharge and it's getting better.” |
| 37 | Standard of care-Future | “Bottom line or the solution to that would probably be to say, “Okay, where are they referred to next? Who's managing that? Who's looking at that?” ” |
| 38 | E-KSB-Barriers | “So we don't have the time in clinical practice to deal with patients individually. And then I think the prescription is issued quickly and then, unfortunately, we have a mishap.” |
| 39 | E-Barriers | “The problem is capacity. We are also a little short of staff,” |
| 40 | E-Barriers | “That's the case with chronic pain patients. The need for us to get people pain-free is very overwhelming, which often leads to changes in treatment over the weekend because people then complain of pain to a new, unfamiliar team. We are less reliable there.” |

| | | |
|---|---|---|
| 41 | Areas for improvement | “And where we're also not so sure is with adverse effects and if there's increasing doses needed to control the pain, there's a complete uncertainty from my perspective about how to deal with that.” |
| 42 | Areas for improvement | “And that's also the danger that these medications will be continued because one thinks time after time - sometimes a different doctor is called in - that it is now good, it's chronic, it's difficult, what do we want to do about it?” |
| 43 | Areas for improvement | “I agree with the argument that they need to be mobilized. And then he has to be pain-compensated, so that's always okay, where does it go and how do you get them out? Does there need to be more inpatient time before the patient has the feeling that they need to go, but they are not compensated? Then you also risk compliance again. It's not easy.” |
| 44 | General practitioner-Statusquo | “But I also think there's a risk that we'll just quickly sign the prescription because the physician assistant will come and request it. And for me, there's also a bit of a potential risk that these things could continue.” |
| 45 | General practitioner-Statusquo | “What I hear from my patients is that they have difficulty finding a GP in the first place because they are no longer accepting new patients, because they don't even have the time” |
| 46 | General practitioner-Future | “And just then it would require instructions to reduce so and so much in such and such weeks. If not, please refer him to a specialist, pain therapist, whatever. Maybe that would help them.” |
| 47 | General practitioner-Future | “Yes, not just the patients, the GPs would receive it too in the end. That would also support the GPs” |
| 48 | Clinician responsibility | “The doctor has to see the patient to check whether there is a wound infection. For example, is there something... Do they suspect something is wrong with the prosthesis? Do you have to look at it again so that you don't miss anything? Or the patient has another problem? Do they perhaps have an infection on top of that, which makes the pain feel stronger again, so that they say: “Okay, then they just need a little more time, they have acute pneumonia, we won't tap them for four days and then we'll go on."” |
| 49 | Clinician responsibility | “If it can't be helped, the patient simply has to contact their GP, for example, and follow up: ”Okay, what is your individual problem? Do you need an examination?” Is there something wrong or is there something that needs to be addressed individually?” |
| 50 | Clinician responsibility | “But yes, I do think that we have a responsibility when we write that on the prescription. At the end of the day.” |

| | | |
|---|---|---|
| 51 | Clinician responsibility | “That's why it should be the attending physician again, basically. I like the idea of the family doctor, the model that he or she overlooks this as a gatekeeper, so to speak.” |
| 52 | Communication | “Yes, if the GP continues this (the opioid therapy) for six weeks and says: “Yes, wait for the follow-up in six weeks.” That's fine. But sometimes we don't get the information for that either.” |
| 53 | P-Barriers | “And I think that's the issue that's not working so well at the moment, because they all go to the GP, the GP doesn't have enough time and then the treatment goes on forever and then at some point they end up with us and then it becomes difficult.” |
| 54 | P-Barriers | “You can't dispense or order it by the tablet at the moment, can you? That isn't possible, is it? That's the problem. You're tied to the pharmaceutical companies, how many tablets they squeeze into the package.” |
| 55 | P-Barriers | “And it's just that, you only get opioids in 30 packs. There is nothing. For NSAIDs, you have five, six days, three days. Depending on the pack size, there are all variants, for the opioids there is actually only one.” |
| 56 | P-Barriers | “I also have to say from my experience that if it's too complicated, such as the second scheme (by Genard et al), I've already found that it takes me a while to understand where I stand as a doctor. It shouldn't be too complicated.” |
| 57 | Interdisciplinarity | “But again, you need someone who can adapt and convert the scheme.” |

### 2. Python script of the opioid reduction calculator

#### 2.1 Interface

«Placeholder»
Please contact the corresponding author for a screenshot of the calculator user interface in German language.

**Figure 2** The interface of the opioid reduction calculator allows to enter the opioid analgesic used for current pain control, a back-up opioid, and the deprescribing target dose. To create a reduction schedule, the calculator computes the schedule based on patient factors whether the patient is discharged, still hospitalized, opioid-naïve or a chronic user.

#### 2.2 Tool architecture

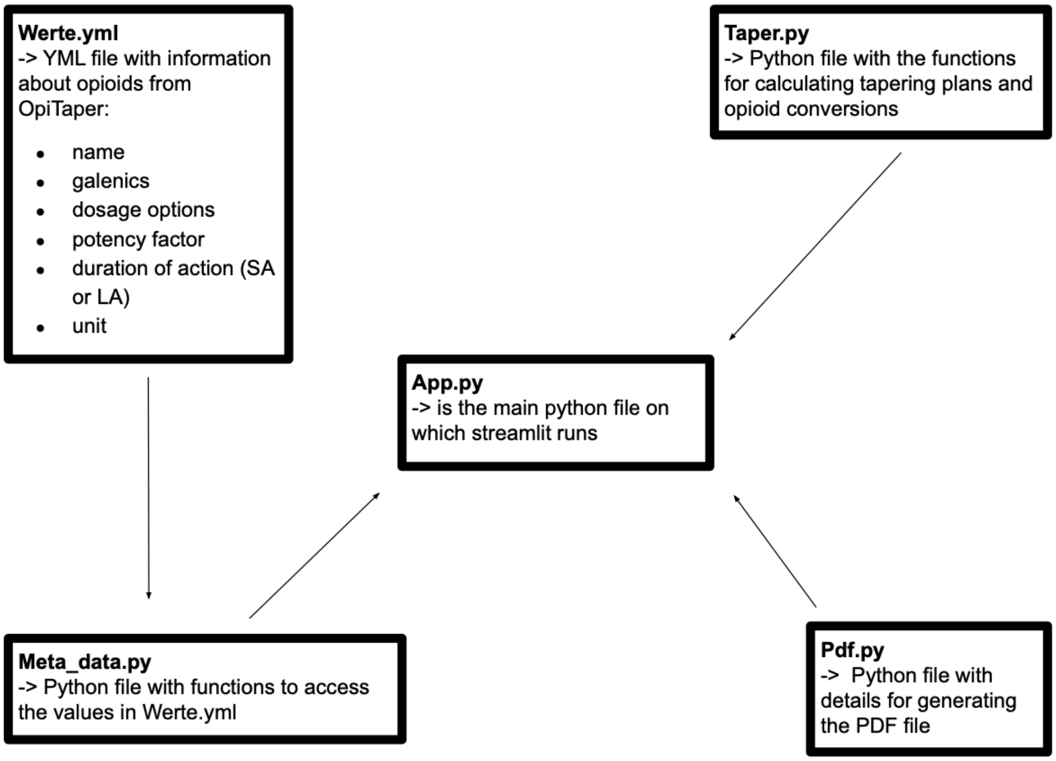

**Figure 3** The structure of OpiTaper includes key Python and YML files. Werte.yml contains detailed opioid information. Meta_data.py accesses Werte.yml, Taper.py calculates tapering schedules and opioid conversions, App.py runs the Streamlit application, and Pdf.py generates PDF tapering plans.

#### 2.3 Code documentation

App.py

```
import os
import streamlit as st
import streamlit_authenticator as stauth
import pandas as pd
from meta_data import get_med_names, get_med, get_same_api_names_SA
from taper import (
generate_plan,
calculate_ome_1,
calculate_original_dose_1,
aequipotenz_berechnen,
)
38from pdf import create_pdf_aqui, create_pdf
import yaml
from yaml.loader import SafeLoader
# Streamlit Benutzeroberfläche
st.set_page_config(
page_title="OpiTaper",
menu_items={
"About": "#Kontaktperson: Marcel Rainer",
```

```
},
)
# Login daten
with open('./config.yml') as file:
config = yaml.load(file, Loader=SafeLoader)
user_name = os.getenv('USER_NAME'
,
config['credentials']['usernames']['ksb']['name'])→
user_password = os.getenv('USER_PASSWORD'
,
config['credentials']['usernames']['ksb']['password'])→
config['credentials']['usernames']['ksb']['name'] = user_name
config['credentials']['usernames']['ksb']['password'] = user_password
authenticator = stauth.Authenticate(
config['credentials'],
config['cookie']['name'],
config['cookie']['key'],
config['cookie']['expiry_days'],
config['pre-authorized']
)
# Login Fenster
authenticator.login()
if st.session_state["authentication_status"]:
authenticator.logout()
st.title("OpiTaper")
all_med_names = get_med_names()
# Aktuelle Medikationen Eingabe
with st.container(border=True):
st.subheader("Aktuelle Medikation")
col1, col2 = st.columns(2)
39current_opioid_name = col1.selectbox(
"Aktuelle Opioid-Fixmedikation:", all_med_names
)
med1 = get_med(current_opioid_name)
unit1 = med1["unit"]
current_opioid_dose = col2.number_input(
"Tagesdosis in mg bzw. µg bei Fentanyl:", min_value=0.0001,
step=0.1
→
)
opioid_same_api_SA = get_same_api_names_SA(current_opioid_name)
if current_opioid_name == "Fentanyl_Effentora (Pflaster)":
current_reserve = col1.selectbox(
"Eingenommene Opioid-Reservemedikation:", all_med_names
)
reserve1 = get_med(current_reserve)
else:
current_reserve = col1.selectbox(
"Eingenommene Opioid-Reservemedikation:",
opioid_same_api_SA→
)
current_reserve_dose = col2.number_input(
"Reservetagesdosis in mg bzw. µg bei Fentanyl:",
min_value=0.0, step=0.1
→
)
target_dose = st.number_input(
"Zielwert der zu erreichenden Tagesdosierung in mg bzw. µg
bei Fentanyl:",
→
min_value=0.0,
step=0.1,
)
if current_reserve_dose > 0.0:
```

```
if current_opioid_name == "Fentanyl_Effentora (Pflaster)":
current_reserve_dose_0 = calculate_ome_1(reserve1,
current_reserve_dose)→
current_reserve_dose = calculate_original_dose_1(
med1, current_reserve_dose_0
)
total_dose = current_opioid_dose + current_reserve_dose
total_dose = current_opioid_dose + current_reserve_dose
st.success(f"{current_opioid_name}: Aktuelle Tagesdosis =
{total_dose} {unit1}")→
# Ausschleich Medikation Einagbe
with st.container(border=True):
st.subheader("Ausschleich Medikation")
col3, col4 = st.columns(2)
40with col3:
taper_med_name = col3.selectbox(
"Opioid für die Erstellung des Ausschleichplans:",
all_med_names→
)
opioid_same_api_SA = get_same_api_names_SA(taper_med_name)
with col4:
taper_reserve = col4.selectbox(
"Reserveopioid für die Erstellung des Ausschleichplans:",
opioid_same_api_SA→
)
option_taper_types = [
"Opioid-naiv stationär",
"Opioid-exposed stationär",
"Opioid-naiv ambulant",
"Opioid-exposed ambulant",
]
schedule_type = st.radio("Typ des Ausschleichplans:",
option_taper_types)→
med = get_med(taper_med_name)
med3 = get_same_api_names_SA(
taper_med_name
) # unten für Reserveausschleichplan können dann nur Opioide mit
gleichen API und SA angewählt werden→
unit = med["unit"]
# Speichern in st.session state, Variabeln bleiben über
→
verschiedene Interaktionen erhalten (zu Beginn auf Null gesetzt,
für Sicherstellung definierter Anfangswert)
→
if "plan_a" not in st.session_state:
st.session_state["plan_a"] = pd.DataFrame()
if "plan_a1" not in st.session_state:
st.session_state["plan_a1"] = pd.DataFrame()
if "plan_a2" not in st.session_state:
st.session_state["plan_a2"] = pd.DataFrame()
if "plan_a3" not in st.session_state:
st.session_state["plan_a3"] = pd.DataFrame()
if "summary" not in st.session_state:
st.session_state["summary"] = pd.DataFrame()
if "plan_b" not in st.session_state:
st.session_state["plan_b"] = pd.DataFrame()
if "REDUCTION_RATE" not in st.session_state:
st.session_state["REDUCTION_RATE"] = 0
41if "REDUCTION_INTERVALL" not in st.session_state:
st.session_state["REDUCTION_INTERVALL"] = 0
if "REDUCTION_START" not in st.session_state:
st.session_state["REDUCTION_START"] = 0
if "last" not in st.session_state:
st.session_state["last"] = 0
```

```
if "y30" not in st.session_state:
st.session_state["y30"] = 0
if "y50" not in st.session_state:
st.session_state["y50"] = 0
if "y100" not in st.session_state:
st.session_state["y100"] = 0
# Auswahl der verschiedenen Reduktionspläne
if st.button("Ausschleichplan erstellen"):
if schedule_type == "Opioid-naiv stationär":
REDUCTION_RATE = 0.25 # 25%
REDUCTION_INTERVALL = 2 # Alle 2 Tage
REDUCTION_START = 0 # Start an Tag 0
if schedule_type == "Opioid-exposed stationär":
REDUCTION_RATE = 0.1 # 10%
REDUCTION_INTERVALL = 3 # Alle 3 Tage
REDUCTION_START = 0 # Start an Tag 0
if schedule_type == "Opioid-naiv ambulant":
REDUCTION_RATE = 0.25 # 25%
REDUCTION_INTERVALL = 2 # Alle 2 Tage
REDUCTION_START = 2 # Start an Tag 2
if schedule_type == "Opioid-exposed ambulant":
REDUCTION_RATE = 0.1 # 10%
REDUCTION_INTERVALL = 3 # Alle 3 Tage
REDUCTION_START = 2 # Start an Tag 2
# Falls aktuelle Medikation ungleich Ausschleichmedikation -> 3
Pläne mit verschiedenen Äquipotenzen werden erstellt→
if med1["conversion"] != med["conversion"]:
try:
(
st.session_state["y100"],
st.session_state["y50"],
st.session_state["y30"],
) = aequipotenz_berechnen(total_dose, med1, med)
(
st.session_state["plan_a1"],
st.session_state["last"],
st.session_state["summary"],
42) = generate_plan(
REDUCTION_RATE,
REDUCTION_INTERVALL,
REDUCTION_START,
st.session_state["y30"],
target_dose,
med,
)
(
st.session_state["plan_a2"],
st.session_state["last"],
st.session_state["summary"],
) = generate_plan(
REDUCTION_RATE,
REDUCTION_INTERVALL,
REDUCTION_START,
st.session_state["y50"],
target_dose,
med,
)
(
st.session_state["plan_a3"],
st.session_state["last"],
st.session_state["summary"],
) = generate_plan(
REDUCTION_RATE,
```

```
REDUCTION_INTERVALL,
REDUCTION_START,
st.session_state["y100"],
target_dose,
med,
)
except ZeroDivisionError:
st.error(
f"Wert {st.session_state['y30']} zu klein! Bitte
korrekten Wert eingeben"→
)
except ValueError:
st.error("Zu kleine Wertdifferenzen")
else:
try:
(
st.session_state["plan_a"],
st.session_state["last"],
st.session_state["summary"],
) = generate_plan(
REDUCTION_RATE,
REDUCTION_INTERVALL,
43REDUCTION_START,
total_dose,
target_dose,
med,
)
st.subheader(f"Ausschleichplan mit {taper_med_name}")
except ZeroDivisionError:
st.error(f"Wert {total_dose} zu klein! Bitte korrekten
Wert eingeben")→
except ValueError:
st.error("Zu kleine Wertdifferenzen")
if taper_med_name == "Fentanyl_Effentora (Pflaster)":
st.write(
"Die angebenen Dosierungen entsprechen der Dosierung pro
→
24h! Dosierungen von 144 g/24h bzw. 72 g/24h
g/24h Pflasters."
→
entsprechen der Hälfte bzw. dem Viertel eines 288
→
)
if (
med1["conversion"] != med["conversion"]
): # dass die 3 Pläne gezeigt werden, wenn aktuelle Medikation
ungleich Ausschleichmedikation→
st.write(f"Mit 30% Äquipotenz, Startwert
{st.session_state['y30'] } {unit}")→
st.dataframe(
st.session_state["plan_a1"],
hide_index=True,
)
st.write(f"Mit 50% Äquipotenz, Startwert
{st.session_state['y50'] } {unit}")→
st.dataframe(
st.session_state["plan_a2"],
hide_index=True,
)
st.write(f"Mit 100% Äquipotenz, Startwert
{st.session_state['y100'] } {unit}")→
st.dataframe(
st.session_state["plan_a3"],
hide_index=True,)
```

```
st.subheader(f"Zusammenfassung Ausschleichplan mit
{taper_med_name} mit 100% Äquipotenz")→
st.dataframe(
st.session_state["summary"],
hide_index=True,
)
else:
44st.dataframe(
st.session_state["plan_a"],
hide_index=True,
)
st.subheader(f"Zusammenfassung Ausschleichplan mit
{taper_med_name}")→
st.dataframe(
st.session_state["summary"],
hide_index=True,
)
if st.button("PDF generieren für Ausschleichplan"):
try:
if (
med1["conversion"] != med["conversion"]
):
pdf_content = create_pdf_aqui(
st.session_state["plan_a1"],
st.session_state["plan_a2"],
st.session_state["plan_a3"],
st.session_state["summary"],
med["name"]
)
else:
pdf_content = create_pdf(
st.session_state["plan_a"],
st.session_state["summary"],
med["name"]
)
except Exception as err:
st.error(f"Error: {err}")
st.download_button(
label="Download PDF für Ausschleichplan",
data=pdf_content,
file_name="Ausschleichplan.pdf",
mime="application/pdf",
)
# Eingabe für Reserve-Ausschleichplan
with st.container(border=True):
st.subheader(f"Reserve Ausschleich Medikation")
planb = st.selectbox("Opioid für einen Reserve-Ausschleichplan:",
med3)→
schedule_type2 = st.radio("Typ des Reserve-Ausschleichplans:",
option_taper_types)→
45st.session_state["last"] = st.number_input(
"Startwert des Reserve-Ausschleichplanes in mg bzw. µg bei
Fentanyl:",
→
min_value=0.0,
step=0.001,
)
med_reserve = get_med(planb)
if st.button("Reserve-Ausschleichplan erstellen"):
if schedule_type2 == "Opioid-naiv stationär":
REDUCTION_RATE = 0.25 # 25%
REDUCTION_INTERVALL = 2 # Alle 2 Tage
REDUCTION_START = 0 # Start an Tag 0
```

```
if schedule_type2 == "Opioid-exposed stationär":
REDUCTION_RATE = 0.1 # 25%
REDUCTION_INTERVALL = 3 # Alle 3 Tage
REDUCTION_START = 0 # Start an Tag 0
if schedule_type2 == "Opioid-naiv ambulant":
REDUCTION_RATE = 0.25 # 25%
REDUCTION_INTERVALL = 2 # Alle 2 Tage
REDUCTION_START = 2 # Start an Tag 2
if schedule_type2 == "Opioid-exposed ambulant":
REDUCTION_RATE = 0.1 # 10%
REDUCTION_INTERVALL = 3 # Alle 3 Tage
REDUCTION_START = 2 # Start an Tag 2
try:
st.session_state["plan_b"], last2,
st.session_state["summary"] = generate_plan(→
REDUCTION_RATE,
REDUCTION_INTERVALL,
REDUCTION_START,
st.session_state["last"],
0,
med_reserve,
)
st.subheader(f"Reserveausschleichplan mit {planb}")
st.dataframe(
st.session_state["plan_b"],
hide_index=True,
)
46st.subheader(f"Zusammenfassung Reserveausschleichplan mit
{planb}")→
st.dataframe(
st.session_state["summary"],
hide_index=True,
)
except ZeroDivisionError:
st.error(
f"Wert {st.session_state['last']} zu klein! Bitte
korrekten Wert eingeben"→
)
except ValueError:
st.error("Zu kleine Wertdifferenzen")
if "plan_b" in st.session_state:
if st.button("PDF generieren Reserve-Ausschleichplan"):
try:
pdf_content = create_pdf(
st.session_state["plan_b"],
st.session_state["summary"],
planb,
)
except Exception as err:
st.error(f"Error: {err}")
st.download_button(
label="Download PDF Reserve-Ausschleichplan",
data=pdf_content,
file_name="Ausschleichplan.pdf",
mime="application/pdf",
)
elif st.session_state["authentication_status"] is False:
st.error('Username/password is incorrect')
elif st.session_state["authentication_status"] is None:
st.warning('Please enter your username and password')
Taper.py
import pandas as pd
from math import ceil
```

```
from functools import partial
import numpy as np
from typing import Union
47# wird genutzt für Reserveberechnung, die Werte, werden gerundet auf
0.5 genau→
def calculate_and_format(value):
first = round(value * 1 / 10 * 2) / 2
second = round(value * 1 / 6 * 2) / 2
return f"{first}- {second}"
"""""→
""" ""

def closest_val_tabletts(val: float, med: dict) -> Union[float, float,
float]:→
values = np.sort(np.array(med["werte"]))
dose = np.array(val)
values_in_dose = dose / values
indices = np.where(values_in_dose >= 2)[0]
if len(indices) <= 1:
multiple = values_in_dose[0].round()
base_val = values[0]
else:
values = values[indices]
tmp = np.array(val) / np.sort(np.array(values))
x = abs(tmp.round() * np.array(values) - val)
arg = np.where(x == x.min())
arg_min = np.argwhere(tmp == min(tmp[arg]))
base_val = values[arg_min[0][0]]
multiple = tmp[arg_min[0][0]].round()
if multiple == 0:
base_val = 0
dose_val = base_val * multiple
return dose_val, multiple, base_val
48# Bestimmt den nächstgelegenen Dosiswert in Tropfenform basierend auf
der Eingabedosis→
def closest_val_drops(val: float, med: dict) -> Union[float, float,
float]:→
base_val = med["werte"][0]
multiple = np.round(val / base_val) # = Anzahl Tropfen
dose_val = multiple * base_val # = Anzahl mg
return dose_val, multiple, base_val
# Bestimmt den nächstgelegenen Dosiswert in Millilitern basierend auf
der Eingabedosis→
def closest_val_ml(val: float, med: dict) -> Union[float, float, float]:
base_val = med["werte"][0]
multiple = np.round(val / base_val)
ml = multiple * med["mass"] # Anzahl Mililiter
dose_val = multiple * base_val # Anzahl mg
return dose_val, ml, base_val
# Bestimmt den nächstgelegenen Dosiswert in Form eines transdermalen
Pflasters basierend auf der Eingabedosis→
def closest_val_patch(val: float, med: dict) -> Union[float, float,
float]:→
tmp_vals = med["werte"]
tmp_vals.append(0) # hier sind kiene Vielfachen zugelassen
values = np.array(tmp_vals)
base_val = values[np.argmin(abs(values - val))] # Anzahl ug
return base_val, 1, base_val
# Bestimmt den nächstgelegenen Dosiswert basierend auf dem
Medikationstyp→
def closest_val(val: float, med: dict) -> Union[float, float, float]:
if med["typ"] == "Tabl" or med["typ"] == "Kaps":
return closest_val_tabletts(val, med)
elif med["typ"] == "Tropf" and med["mass_einheit"] == "tropfen":
```

```
return closest_val_drops(val, med)
elif med["typ"] == "Tropf" and med["mass_einheit"] == "ml":
return closest_val_ml(val, med)
elif med["typ"] == "Transdermal":
return closest_val_patch(val, med)
# Funktion zur Berechnung der OME (Orale Morphinäquivalente) von
Nicht-Fentanyl für Rechnung am Anfang→
def calculate_ome_1(opioid, dose):
ome_dose = dose * opioid["conversion"]
49return ome_dose
# Funktion zur Rückberechnung von OME zur ursprünglichen
→
Opioid-Dosierung von nicht Fentanyl zu Fentanyl für Rechnung am
Anfang
→
def calculate_original_dose_1(opioid, ome):
original = round(ome * 1000 * (1 / opioid["conversion"]))
return original
# Funktion zur Berechnung der OME (Orale Morphinäquivalente)
def calculate_ome(opioid: dict, dose: float) -> float:
if opioid["unit"] == "ug":
dose = dose / 1000
ome_dose = dose * opioid["conversion"]
return ome_dose
# Funktion zur Rückberechnung von OME zur ursprünglichen
Opioid-Dosierung→
def calculate_original_dose(opioid, ome):
original = round(ome * (1 / opioid["conversion"]))
if opioid["unit"] == "ug":
original = original * 1000
return original
# Berechnung verschiedene äquipotenzen
def aequipotenz_berechnen(dose, von_opioid, zu_opioid):
anfang = calculate_ome(von_opioid, dose)
y100 = calculate_original_dose(zu_opioid, anfang)
y50 = y100 * 0.5
y30 = y100 * 0.3
return y100, y50, y30
def generate_plan(
red_rate, # Reduktionsrate (0.1 oder 0.25)
red_intervall, # Nach wie vielen Tagen der erneute
Reduktionsschritt stattfinden soll→
red_start, # An welchem Tag die erste Reduktion stattfinden soll
start_dose, # Die Startdosierung, die aktuelle Medikation
end_dose, # Bis au welche Dosierung man reduzieren möchte
med,
): # Angabe mit welchem Opioid aus der Liste man reduzieren möchte
if med["name"] in "Fentanyl_Effentora (Pflaster) ":
50red_intervall = 3
reduction = start_dose * red_rate # Berechnet Reduktion pro Schritt
unit = med["unit"] # gibt Einheit des ausgewählten Opioids an
steps = ceil((start_dose - end_dose) / reduction) - 1 # Berechnet
Anzahl Reduktionsschritte
→
doses = [max(start_dose - i * reduction, end_dose) for i in
→
range(1,steps + 1)] # berechnet verschiedene Dosierungen und
schaut, dass nicht unter end_dose
→
doses = [dose for dose in doses if dose >= end_dose and dose > 0]
duplicated_list = [dose for dose in doses for
in
_
range(red_intervall)]→
days = list(range(1, len(duplicated_list) + red_start + 1)) #
```

```
Berechnet die totale Anzahl Ausschleichstage→
duplicated_list = [
x for x in doses for
_
in range(red_intervall)
] # Erweitert jede Dosierung für jeden Tag
for
_
in range(red_start):
duplicated_list.insert(0, start_dose)
# Erstellt df, damit der Plan dargestellt werden kann
dose_col = f"Dosis ({unit})"
df = pd.DataFrame(
{
"Tag": days,
f"Dosis ({unit})": duplicated_list,
}
)
df[f"Reserve ({unit})"] = df[dose_col].apply(calculate_and_format)
fixed_closest_value = partial(closest_val, med=med) #unterschiedliche
Dartstellung des Planes, je nach Verabreichungsform→
if med["typ"] == "Tabl" or med["typ"] == "Kaps":
df[f"Nächste Dosis ({unit})"], df["Anzahl Tabl/Kaps"],
df["Basiswert"] = zip(→
*df[dose_col].apply(fixed_closest_value)
)
last_value = df[f"Nächste Dosis ({unit})"].iloc[-1]
elif med["typ"] == "Tropf" and med["mass_einheit"] == "tropfen":
df[f"Nächste Dosis ({unit})"], df["Anzahl Tropfen"],
df["Basiswert"] = zip(→
*df[dose_col].apply(fixed_closest_value)
)
last_value = df[f"Nächste Dosis ({unit})"].iloc[-1]
elif med["typ"] == "Tropf" and med["mass_einheit"] == "ml":
df[f"Nächste Dosis ({unit})"], df["Total Milliliter"],
df["Basiswert"] = zip(→
*df[dose_col].apply(fixed_closest_value)
)
51last_value = df[f"Nächste Dosis ({unit})"].iloc[-1]
elif med["typ"] == "Transdermal":
df[f"Nächste Dosis ({unit})"], df["Anzahl Pflaster"],
df["Basiswert"] = zip(→
*df[dose_col].apply(fixed_closest_value)
)
last_value = df[f"Nächste Dosis ({unit})"].iloc[-1]
if med["name"] in "Fentanyl_Effentora (Pflaster) ": #Spezialfall
Fentanyl wegen "" und red_intervall immer 3 Tage→
red_intervall = 3
columns_to_replace = [
f"Nächste Dosis ({unit})",
dose_col,
"Anzahl Pflaster",
"Basiswert",
]
num_rows = len(df)
for i in range(0, num_rows, 3):
start_index = i
# In erster Reihe Wert anzeigen
df.loc[start_index, columns_to_replace] = df.loc[
start_index, columns_to_replace
]
# Die nächsten 2 Reihen mit "-" ersetzen
if start_index + 1 < num_rows:
```

```
df.loc[start_index + 1, columns_to_replace] = "-"
if start_index + 2 < num_rows:
df.loc[start_index + 2, columns_to_replace] = "-"
# Erstellt eine gefilterte Version von df ohne die "-" für die
Zusammenfassung→
summary_df_filtered = df[df["Basiswert"] != "-"]
else:
# Konvertiere die Dosis-Spalte in numerische Werte und filtere
Nullwerte heraus
→
df[dose_col] = pd.to_numeric(df[dose_col], errors='coerce')
df = df[df[dose_col] > 0].reset_index(drop=True)
# Zusammenfassung Dataframes -> anzeigen Anzahl Tabl/Tropfen etc
und für welche Stärke→
if med["typ"] == "Tabl" or med["typ"] == "Kaps":
summary_df = df.groupby("Basiswert")["Anzahl
Tabl/Kaps"].sum().reset_index()→
summary_df.columns = ["Basiswert", "Total Anzahl Tabl/Kaps"]
elif med["typ"] == "Tropf" and med["mass_einheit"] == "tropfen":
52summary_df = df.groupby("Basiswert")["Anzahl
Tropfen"].sum().reset_index()→
summary_df.columns = ["Basiswert", "Total Anzahl Tropfen"]
elif med["typ"] == "Tropf" and med["mass_einheit"] == "ml":
summary_df = df.groupby("Basiswert")["Total
Milliliter"].sum().reset_index()→
summary_df.columns = ["Basiswert", "Total Milliliter"]
elif med["typ"] == "Transdermal":
summary_df = summary_df_filtered.groupby("Basiswert")["Anzahl
Pflaster"].sum().reset_index()→
summary_df.columns = ["Basiswert", "Total Anzahl Pflaster"]
return df, last_value, summary_df
```

Meta_data.py

```
import yaml
# Funktion um YAML-Datei zu lesen
def read_yaml(file_path):
with open(file_path, "r") as file:
return yaml.safe_load(file)
data = read_yaml("werte.yml")
def get_med_names() -> list:
"""Zugriff auf die Medikamentennamen aus Werte.yml
Returns:
_type_: Namen der Medikamente
"""
tmp = [n["name"] for n in data["medikamente"]]
return tmp
def get_med(opioid_name: str) -> dict:
"""Zugriff Medikamenten Daten aus Werte.yml
Args:
name (str): Name des Medikamentes
Returns:
dict: Alle Werte des eigegebenen Medikamentes"""
try:
53med = next(med for med in data["medikamente"] if med["name"] ==
opioid_name)→
except Exception as ex:
print(ex)
med = {"name": None}
return med
def get_same_api_names_SA(opioid_name: str) -> list:
"""Erhalt Medikamente mit gleichem Wirkstoff wie opioid_name und
sie müssen SA sein
→
Args:
opioid_name (str): Eingabe des Medikamentes von dem man
```

```
Medikamente mit gleichem Wirkstoff möchte→
Returns:
sind SA
→
list: Medikamente mit gleichem Wirkstoff wie opioid_name und
"""
med_conversion = get_med(opioid_name)["conversion"]
tmp = [
n["name"]
for n in data["medikamente"]
if n["conversion"] == med_conversion and n["wirksamkeit"] == "SA"
return tmp
]
Pdf.py
from fpdf import FPDF
import pandas as pd
# Funktionen, um PDF zu generieren, rein inhaltlich wird da nichts
geändert, geht nur um Layout und was dargestellt werden soll→
def create_pdf(schedule:pd.DataFrame,
summary:pd.DataFrame,
opioid:str,
schedule_reserve:pd.DataFrame=pd.DataFrame(),
)->bytes:
pdf = FPDF()
pdf.add_page()
pdf.set_font("Arial", size=14)
pdf.cell(200, 10, txt="Opioid Tapering Tool", ln=True, align="C")
pdf.set_font("Arial", size=12)
54pdf.cell(200, 10, txt=f"Ausschleichplan für {opioid}", ln=True,
align="C")→
pdf.ln(10)
# Create table header
pdf.set_font("Arial", size=10, style="B")
for i, col in enumerate(schedule.columns):
if i == 0: # Die erste Spalte
pdf.cell(10, 10, col, 1, 0, "C") # Verkleinere die erste
Spalte→
else:
pdf.cell(35, 10, col, 1, 0, "C")
pdf.ln()
pdf.set_font("Arial", size=10)
# Insert table data
for index, row in schedule.iterrows():
for i, col in enumerate(schedule.columns):
if i == 0: # Die erste Spalte
pdf.cell(10, 10, str(row[col]), 1, 0, "C") # Verkleinere
die erste Spalte→
else:
pdf.cell(35, 10, str(row[col]), 1, 0, "C")
pdf.ln()
pdf.ln(10)
# Create Reserve Plan table if input is provided
if not schedule_reserve.empty:
pdf.set_font("Arial", size=12, style="B")
pdf.cell(200, 10, txt="Reserveplan", ln=True, align="L")
pdf.set_font("Arial", size=10, style="B")
for col in schedule_reserve.columns:
pdf.cell(35, 10, col, 1, 0, "C")
pdf.ln()
pdf.set_font("Arial", size=10)
for index, row in schedule_reserve.iterrows():
for col in schedule_reserve.columns:
pdf.cell(35, 10, str(row[col]), 1, 0, "C")
```

```
pdf.ln()
pdf.ln(10)
55# Add summary table
pdf.set_font("Arial", size=12, style="B")
pdf.cell(200, 10, txt="Zusammenfassung", ln=True, align="L")
pdf.set_font("Arial", size=10, style="B")
for col in summary.columns:
pdf.cell(50, 10, col, 1, 0, "C")
pdf.ln()
pdf.set_font("Arial", size=10)
for index, row in summary.iterrows():
for col in summary.columns:
pdf.cell(50, 10, str(row[col]), 1, 0, "C")
pdf.ln()
pdf.ln(10)
pdf.set_font("Arial", size=10)
return pdf.output(dest="S").encode("latin1")
def table_format(schedule, pdf):
# Create table header
pdf.set_font("Arial", size=10, style="B")
for i, col in enumerate(schedule.columns):
if i == 0: # Die erste Spalte
pdf.cell(10, 10, col, 1, 0, "C") # Verkleinere die erste
Spalte→
else:
pdf.cell(35, 10, col, 1, 0, "C")
pdf.ln()
pdf.set_font("Arial", size=10)
# Insert table data
for index, row in schedule.iterrows():
for i, col in enumerate(schedule.columns):
if i == 0: # Die erste Spalte
pdf.cell(10, 10, str(row[col]), 1, 0, "C") # Verkleinere
die erste Spalte→
else:
pdf.cell(35, 10, str(row[col]), 1, 0, "C")
pdf.ln()
pdf.ln(10)
def create_pdf_aqui(schedule1:pd.DataFrame,
56schedule2:pd.DataFrame,
schedule3:pd.DataFrame,
summary:pd.DataFrame,
opioid:str,
schedule_reserve:pd.DataFrame=pd.DataFrame(),
)->bytes:
pdf = FPDF()
pdf.add_page()
pdf.set_font("Arial", size=14)
pdf.cell(200, 10, txt="Opioid Tapering Tool", ln=True, align="C")
pdf.set_font("Arial", size=12)
pdf.cell(200, 10, txt=f"Ausschleichplan für {opioid}", ln=True,
align="C")→
pdf.ln(10)
# Create table header
pdf.set_font("Arial", size=12, style="B")
pdf.cell(200, 10, txt="Äquipotenz 30%", ln=True, align="L")
table_format(schedule1, pdf)
pdf.set_font("Arial", size=12, style="B")
pdf.cell(200, 10, txt="Äquipotenz 50%", ln=True, align="L")
table_format(schedule2, pdf)
pdf.set_font("Arial", size=12, style="B")
pdf.cell(200, 10, txt="Äquipotenz 100%", ln=True, align="L")
table_format(schedule3, pdf)
```

```
# Create Reserve Plan table if input is provided
if not schedule_reserve.empty:
pdf.set_font("Arial", size=12, style="B")
pdf.cell(200, 10, txt="Reserveplan", ln=True, align="L")
pdf.set_font("Arial", size=10, style="B")
for col in schedule_reserve.columns:
pdf.cell(35, 10, col, 1, 0, "C")
pdf.ln()
pdf.set_font("Arial", size=10)
for index, row in schedule_reserve.iterrows():
for col in schedule_reserve.columns:
pdf.cell(35, 10, str(row[col]), 1, 0, "C")
pdf.ln()
pdf.ln(10)
57# Add summary table
pdf.set_font("Arial", size=12, style="B")
pdf.cell(200, 10, txt="Zusammenfassung", ln=True, align="L")
pdf.set_font("Arial", size=10, style="B")
for col in summary.columns:
pdf.cell(50, 10, col, 1, 0, "C")
pdf.ln()
pdf.set_font("Arial", size=10)
for index, row in summary.iterrows():
for col in summary.columns:
pdf.cell(50, 10, str(row[col]), 1, 0, "C")
pdf.ln()
pdf.ln(10)
pdf.set_font("Arial", size=10)
return pdf.output(dest="S").encode("latin1")
def create_pdf_schedule(schedule, opioid):
pdf = FPDF()
pdf.add_page()
pdf.set_font("Arial", size=14)
pdf.cell(200, 10, txt="Opioid Tapering Tool", ln=True, align="C")
pdf.set_font("Arial", size=12)
pdf.cell(200, 10, txt=f"Ausschleichplan für {opioid}", ln=True,
align="C")→
pdf.ln(10)
# Create table header
pdf.set_font("Arial", size=10, style="B")
for i, col in enumerate(schedule.columns):
if i == 0: # Die erste Spalte
pdf.cell(10, 10, col, 1, 0, "C") # Verkleinere die erste
Spalte→
else:
pdf.cell(35, 10, col, 1, 0, "C")
pdf.ln()
58pdf.set_font("Arial", size=10)
# Insert table data
for index, row in schedule.iterrows():
for i, col in enumerate(schedule.columns):
if i == 0: # Die erste Spalte
pdf.cell(10, 10, str(row[col]), 1, 0, "C") # Verkleinere
die erste Spalte→
else:
pdf.cell(35, 10, str(row[col]), 1, 0, "C")
pdf.ln()
pdf.ln(10)
pdf.set_font("Arial", size=10)
return pdf.output(dest="S").encode("latin1")
```

Werte.yml

```
medikamente:
- name: Tramadol_Tramal 100mg/ml (Tropf)
```

typ: Tropf
werte: [2.5]
conversion: 0.15
mass: 1
mass_einheit: "tropfen"
wirksamkeit: SA
unit: "mg"
- name: Tramadol_Tramal Retard (Tabl)
typ: Tabl
werte: [200,150,100,50]
conversion: 0.15
wirksamkeit: LA
unit: "mg"
- name: Morphin 10 mg/ml (Tropf)
typ: Tropf
werte: [0.5]
conversion: 1
mass: 1
mass_einheit: "tropfen"
wirksamkeit: SA
unit: "mg"
59- name: Morphin 20 mg/ml (Tropf)
typ: Tropf
werte: [1.0]
conversion: 1
mass: 1
mass_einheit: "tropfen"
wirksamkeit: SA
unit: "mg"
- name: Morphin_MST Continus Retard (Tabl)
typ: Tabl
werte: [100,60,30,10]
conversion: 1
wirksamkeit: LA
unit: "mg"
- name: Methadon_Ketalgin (Tabl)
typ: Tabl
werte: [40,20,10,5]
conversion: 3
wirksamkeit: LA
unit: "mg"
- name: Oxycodon_Oxynorm 10mg/ml (Tropf)
typ: Tropf
werte: [1.0]
conversion: 2
mass: 0.1
mass_einheit: "ml"
wirksamkeit: SA
unit: "mg"
- name: Oxycodon_Oxycontin Retard (Tabl)
typ: Tabl
werte: [80,40,20,10,5]
conversion: 2
wirksamkeit: LA
unit: "mg"
- name: Oxycodon/Nalaxon_Targin Retard (Tabl)
typ: Tabl
werte: [80,60,40,20,10,5]
conversion: 2
wirksamkeit: LA
unit: "mg"
- name: Hydromorphon_Palladon (Kaps)
typ: Kaps

```
60werte: [2.6,1.3]
conversion: 8
wirksamkeit: SA
unit: "mg"
- name: Hydromorphon_Palladon Retard (Kaps)
typ: Kaps
werte: [24,16,8,4]
conversion: 8
wirksamkeit: LA
unit: "mg"
- name: Fentanyl_Effentora (Pflaster) #werte als Dosis, die
→
innerhalb von 24h verarbreit wird angegeben / letzte 2 Dosierungen
1/2 bzw. 1/2 von letztem Pflaster
→
typ: Transdermal
werte: [2400,1800,1200,900,600,288,144,72]
conversion: 100
wirksamkeit: LA
unit: "ug"
```

### 3. Delphi survey

#### 3.1 Delphi survey consensus items

**Table 3** The findings of the focus group discussion led to the synthesis of 49 recommendations regarding deprescribing determinants and the opioid reduction calculator. These were then validated and revised through a Delphi survey with clinicians from a broader range of medical specialties.

| | Tapering plan |
|---|---|
| | **Reduction rate** |
| 1 | It is essential to taper opioids only in patients who agree to do so. |
| ***1 revised*** | ***It is essential to taper opioids. In patients with chronic pain, agreement should be sought.*** |
| 2 | The tapering rate may need to be adjusted or stopped if withdrawal symptoms appear or pain intensifies. |
| 3 | A reduction rate of opioids should be within 5-25% of the previous daily intake. |
| 4 | Patients with chronic opioid use may be tapered 10% of their previous daily intake. |
| 5 | Patients without chronic opioid use may be tapered 20-25% of their previous daily intake. |
| 6 | When tapering, the daily opioid dose should be reduced every 2-3 days. |
| | **Duration** |
| 7 | After surgery, patients are expected to return to their baseline opioid dose within 6 weeks. |
| 8 | After surgery, tapering opioids for postoperative pain control may take 10 to 14 days. |
| ***8 revised*** | ***The taper length phase can vary postoperatively. Pain control may be tapered sooner for minimally invasive procedures and longer for other procedures.*** |
| | **Initiation of tapering** |

| | |
|---|---|
| 9 | Opioid dose reductions need to be coherent with the level of the patient's mobilisation. |
| 10 | Opioids should not be reduced before patients regain their mobility. |
| ***10 revised*** | ***Tapering may begin before full mobility is restored, but must not interfere with mobility recovery.*** |
| 11 | After discharge of patients, the tapering should start or continue with a 2-day pause before opioids are (further) reduced. |
| | **Adaptibility** |
| 12 | During the tapering process, patients need to have the option of taking back-up opioids. |
| 13 | Back-up opioids should preferably be short-acting and constitute 1/10 to 1/6 of the fixed dose. |
| 14 | To taper off opioid patches, these should be either cut (halves, quarters) or converted to an oral opioid. |
| | **Design** |
| 15 | An automated tapering tool should be easy to use by providing drop-down menus that calculate the respective tapering route. |
| 16 | The pain history (e.g. type of injury) should inform the length of the tapering by reducing the reduction rate, for example. |
| 17 | The tool should allow to define a patient-specific goal to reduce the dose of opioids to. |
| | **Recipient** |
| 18 | The output of the tool needs to be easy to follow for patients. |
| 19 | The output of the tool needs to be easy to follow for the next caring healthcare professional. |
| | **Patient pain education** |
| | **Paper form** |
| 20 | An information sheet needs to address if patients have difficulties following the tapering, the general practitioner or the tapering team should be contacted. |
| 21 | This sheet should also encourage patients to be physically active as much as possible. |

| | |
|---|---|
| | **In person** |
| 22 | To maximize compliance, patient's expectations need to be aligned with functional goals and the tapering process. |
| 23 | Functional goals should entail small steps from taking longer walks to spending more time outside. |
| 24 | People should be educated that pain may persist and back-up opioids may be used during the tapering. |
| 25 | Patients should be instructed to prioritize stopping opioids first, then other analgesics (e.g. paracetamol, metamizol). |
| 26 | Patients need to be informed that tablets are not a substitute for physical activity in the recovery process. |
| 27 | Patients may need to be interviewed if pain levels and functionality do not match. |
| | **Individualised patient care** |
| | **Type of pain** |
| 28 | Opioids in surgical patients with preexisting chronic pain (e.g. arthrosis) should be tapered slower. |
| | **Opioid exposure** |
| 29 | Opioid-naive patients who required high opioid dosages during their hospitalization (e.g. analgosedation with remifentanil on a critical care ward) should also be tapered as if they were non-naive. |
| ***29 revised*** | ***Opioid-naive patients exposed to high doses may taper quickly if exposure was short-term.*** |
| 30 | Ideally, patients with chronic opioid use are tapered below their baseline after surgery. |
| 31 | The last total opioid dose (= used back-up + fixed doses) should inform the starting dose of the tapering. |

| | |
|---|---|
| | **Other clinical parameters** |
| 32 | Renal and hepatic impairment should be considered when choosing an opioid and concomitant analgesia. |
| 33 | Reports of physiotherapy should inform the tapering process. |
| 34 | Patients who catastrophize should be tapered slower. |
| ***34 revised*** | ***Catastrophizing patients may also taper fast if closely monitored for withdrawal symptoms and pain.*** |
| 35 | Other sources of pain need to be identified to avoid cross-prescribing opioids/pain medication (e.g. hematoma, infection). |
| | **Barriers** |
| 36 | NRS scores should be considered with caution due to patient misconceptions. |
| 37 | Only one dose should be prescribed to avoid mixing different packs (e.g. 15 mg and 10 mg). |

| | |
|---|---|
| | **Pain management** |
| | **Postoperative care** |
| 38 | Patients discharged with a higher dose than before admission should be followed up by phone calls while tapering. |
| 39 | Opioids in cancer patients after surgery should also be tapered if they don't experience pain from metastases. |
| 40 | Patients should be prescribed short-acting opioids before long-acting opioids to target pain. This reduces drug side effects at times when an analgesic is not needed. |
| ***40 revised*** | ***Short-acting opioids should be preferred but use of long-acting opioids should not be ruled out.*** |

| | |
|---|---|
| | **Areas for improvement** |
| 41 | Pain medication should be reviewed routinely to avoid continuation without indication. |

| | |
|---|---|
| | **Clinician responsibility** |
| 42 | The primary responsibility for postoperative pain control after discharge lies with the general practitioner. |

*Please open the provided instruction manual of the tool and open the tool to test it.*

| | |
|---|---|
| | **Please rate the success of implementation of the items above** |
| 43 | **Reduction rate** |
| 44 | **Duration** |
| 45 | **Initiation of tapering** |
| 46 | **Adaptibility** |
| 47 | **Design** |
| 48 | **Recipient** |
| 49 | **Paper form** |

#### 3.2 Delphi survey non-consensus items

**Table 4** For six items of the Delphi survey, the predefined consensus level (≥80%) was not reached in the first round. These were rephrased based on comments from survey participants and included in the second round (see section 3.3).

| Segment | Statements without consensus | Agreement rating (1=disagreement, 7=agreement)<br>Mean | 95% CI | Level of consensus |
|---|---|---|---|---|
| **Tapering plan** | It is essential to taper opioids only in patients who agree to do so. | 3.8 | 2.2-5.4 | 50% |
| | Tapering opioids for postoperative pain control may take 10 to 14 days. | 4.4 | 3.1-5.7 | 60% |
| | Opioids should not be reduced before patients regain their mobility. | 3.6 | 2.3-4.9 | 50% |
| **Individualized patient care** | Opioid-naive patients who required high opioid dosages during their hospitalization (e.g. on a critical care ward) should also be tapered slowly. | 4.7 | 3.3-6.1 | 70% |
| | Patients who catastrophize should be tapered slower. | 4.1 | 2.8-5.4 | 60% |
| **Pain management** | Patients should be prescribed short-acting opioids before long-acting opioids to target pain. | 4.6 | 3.3-5.9 | 70% |

#### 3.3 Revision of non-consensus items

***Proposed solutions to missing consensus:***

**Tapering plan**

- It is essential to taper opioids. In patients with chronic pain, agreement should be sought.
- The taper length phase can vary postoperatively. Pain control may be tapered sooner for minimally invasive procedures and longer for other procedures.
- Tapering may begin before full mobility is restored, but must not interfere with mobility recovery.

**Individualized patient care**

- Opioid-naive patients exposed to high doses may taper quickly if exposure was short-term.
- Catastrophizing patients may also taper fast if closely monitored for withdrawal symptoms and pain.

**Pain management**

- Short-acting opioids should be preferred but use of long-acting opioids should not be ruled out.

### 4. Patient interview questions

#### 4.1 Interview Understandability Patient Brochure

This questionnaire is used to document the answers to our questions about the understandability of the sentence structures in the patient brochure. Please do not record any patient characteristics!

**Interviewer**

Approximate time the patient spent reading the brochure.

Please tell me in your own words what this document is about.

Could you please tell me how to dispose of a used transdermal patch?

Another specific comprehension question. Selected by the interviewer.

Are there any sentences that are difficult to understand? Do you have a suggestion for a different wording?

On a scale of 0 to 5, where 0 is not understandable at all and 5 is very understandable: How easy did you find the brochure to understand?

How can we make it even easier to understand?

Were there any sentences that made you anxious?

Are there any sentences that we could omit?

Is there any information you are still missing?

What would you change about the layout?

Would a brochure like this help you learn more about the medicine, how to take it and any problems that may occur?

Would you recommend this brochure to other patients?

Notes from the interview. For statements made by the patient that do not fit elsewhere or were made in a follow-up question.

Observations of the interviewer. Visible uncertainty on the part of the patient, etc.

#### 4.2 Interview Understandability Reduction Plan

This questionnaire is used to document the answers to our questions about the understandability of the redcution plan. Please do not record any patient characteristics!

**Interviewer**

Please tell me in your own words what this document is about.

Could you please tell me how many and which tablets you would take on day X?

Another specific comprehension question. Selected by the interviewer.

Are there any elements or sentences that you do not understand? Do you have any suggestions for different wording?

On a scale of 0 to 5, where 0 is not understandable at all and 5 is very understandable: How understandable did you find this redcution plan?

How can we make the redcution plan even easier to understand?

Is there any information missing?

What would you change about the structure?

Would such a redcution plan help you to stop taking your medication?

Would you recommend this redcution plan to other patients?

Notes from the interview. For statements made by the patient that do not fit elsewhere or were made in a follow-up question.

Observations of the interviewer. Visible uncertainty on the part of the patient, etc.

### 5. Reporting checklists

**Table 5** Consolidated criteria for reporting qualitative studies (COREQ): 32-item checklist[1]

| No | Item | Guide questions/description | Location |
|---|---|---|---|
| **Domain 1: Research team and reflexivity** | | | |
| Personal Characteristics | | | |
| 1. | Interviewer/facilitator | Which author/s conducted the interview or focus group? | Supplement section 1 |
| 2. | Credentials | What were the researcher's credentials? *E.g. PhD, MD* | Supplement section 1 |
| 3. | Occupation | What was their occupation at the time of the study? | Supplement section 1 |
| 4. | Gender | Was the researcher male or female? | Supplement section 1 |

| No | Item | Guide questions/description | Location |
|---|---|---|---|
| 5. | Experience and training | What experience or training did the researcher have? | Supplement |
| Relationship with participants | | | |
| 6. | Relationship established | Was a relationship established prior to study commencement? | Supplement section 1 |
| 7. | Participant knowledge of the interviewer | What did the participants know about the researcher? e.g. *personal goals, reasons for doing the research* | Supplement section 1 |
| 8. | Interviewer characteristics | What characteristics were reported about the interviewer/facilitator? e.g. *Bias, assumptions, reasons and interests in the research topic* | Supplement section 1 |
| **Domain 2: study design** | | | |
| Theoretical framework | | | |

| No | Item | Guide questions/description | Location |
|---|---|---|---|
| 9. | Methodological orientation and Theory | What methodological orientation was stated to underpin the study? *e.g. grounded theory, discourse analysis, ethnography, phenomenology, content analysis* | Methods |
| Participant selection | | | |
| 10. | Sampling | How were participants selected? *e.g. purposive, convenience, consecutive, snowball* | Methods |
| 11. | Method of approach | How were participants approached? e.*g. face-to-face, telephone, mail, email* | Supplement section 1 |
| 12. | Sample size | How many participants were in the study? | Results |
| 13. | Non-participation | How many people refused to participate or dropped out? Reasons? | Supplement section 1 |
| Setting | | | |

| No | Item | Guide questions/description | Location |
|---|---|---|---|
| 14. | Setting of data collection | Where was the data collected? e.g. *home, clinic, workplace* | Methods |
| 15. | Presence of non-participants | Was anyone else present besides the participants and researchers? | Supplement section 1 |
| 16. | Description of sample | What are the important characteristics of the sample? *e.g. demographic data, date* | Results |
| Data collection | | | |
| 17. | Interview guide | Were questions, prompts, guides provided by the authors? Was it pilot tested? | Methods |
| 18. | Repeat interviews | Were repeat interviews carried out? If yes, how many? | Supplement section 1 |
| 19. | Audio/visual recording | Did the research use audio or visual recording to collect the data? | Methods |

| No | Item | Guide questions/description | Location |
|---|---|---|---|
| 20. | Field notes | Were field notes made during and/or after the interview or focus group? | Supplement section 1.1 |
| 21. | Duration | What was the duration of the interviews or focus group? | Methods |
| 22. | Data saturation | Was data saturation discussed? | Supplement section 1, Discussion |
| 23. | Transcripts returned | Were transcripts returned to participants for comment and/or correction? | Results |
| **Domain 3: analysis and findings**z | | | |
| Data analysis | | | |
| 24. | Number of data coders | How many data coders coded the data? | Methods |

| No | Item | Guide questions/description | Location |
|---|---|---|---|
| 25. | Description of the coding tree | Did authors provide a description of the coding tree? | Supplement section 1.2 |
| 26. | Derivation of themes | Were themes identified in advance or derived from the data? | Methods, results |
| 27. | Software | What software, if applicable, was used to manage the data? | Methods |
| 28. | Participant checking | Did participants provide feedback on the findings? | Results |
| Reporting | | | |
| 29. | Quotations presented | Were participant quotations presented to illustrate the themes / findings? Was each quotation identified? e.g. *participant number* | Supplement section 1.4 |
| 30. | Data and findings consistent | Was there consistency between the data presented and the findings? | Results, supplement sections 1.3 and 1.4 |

| No | Item | Guide questions/description | Location |
|---|---|---|---|
| 31. | Clarity of major themes | Were major themes clearly presented in the findings? | Results, figure 2 |
| 32. | Clarity of minor themes | Is there a description of diverse cases or discussion of minor themes? | Results, discussion |

[1] Tong A, Sainsbury P, Craig J. Consolidated criteria for reporting qualitative research (COREQ): a 32-item checklist for interviews and focus groups. Int J Qual Health Care. 2007 Sep 16;19(6):349–57.

**Table 6** A checklist for mixed-methods research manuscript preparation and review[2]

| | |
|---|---|
| Rational and description of MMR design | ☒ Provide a clear statement of the study purpose |
| | ☒ Explicitly describe the MMR design in accordance with Creswell's (2015) typology and use a diagram to illustrate the relationship and sequence of qualitative and quantitative research components |
| | ☒ Justify why the MMR design is appropriate for meeting the study purpose |
| Transparency in describing method details | ☒ Describe the study population(s) and sample(s; e.g., who, what, how many) |
| | ☒ Describe the sampling procedures (including inclusion and exclusion criteria, recruitment) |
| | ☒ Describe qualitative data collection processes (how often data were collected, who collected the data, what kind of data collection instruments were used, how data were recorded—e.g., notes, transcripts) |
| | ☒ Describe quantitative data collection processes (how often data were collected, who collected the data, what kind of data collection instruments were used measurements, validity/reliability) |
| | ☒ Describe qualitative data analysis processes (coding, single or multiple coders, replication logic, credibility) |
| | ☒ Describe quantitative data analysis procedures (missing data and how they are handled, statistical tests used) |
| Integration of qualitative and quantitative research components | ☒ Interpret qualitative analysis results with appropriate quotes if necessary |
| | ☒ Interpret quantitative analysis results in consideration of statistical significance, selection bias, and threats to validity |
| | ☒ Compare qualitative and quantitative results |
| | ☒ Address divergencies and inconsistencies between qualitative and quantitative results |

[2] Lee SYD, Iott B, Banaszak-Holl J, Shih SF, Raj M, Johnson KE, et al. Application of Mixed Methods in Health Services Management Research: A Systematic Review. Med Care Res Rev. 2022 Jun;79(3):331–44.
